# The effectiveness of social bubbles as part of a Covid-19 lockdown exit strategy, a modelling study

**DOI:** 10.1101/2020.06.05.20123448

**Authors:** Trystan Leng, Connor White, Joe Hilton, Adam Kucharski, Lorenzo Pellis, Helena Stage, Nick Davies, CMMID-Covid-19 WG, Matt J. Keeling, Stefan Flasche

**Affiliations:** The Zeeman Institute for Systems Biology & Infectious Disease Epidemiology Research, University of Warwick, UK; Centre for Mathematical Modelling of Infectious Diseases, London School of Hygiene & Tropical Medicine, UK; Department of Mathematics, The University of Manchester, UK

**Keywords:** Contact clustering, Social bubble, Exit strategy, Covid-19

## Abstract

**Background:** During the Covid-19 lockdown, contact clustering in social bubbles may allow extending contacts beyond the household at minimal additional risk and hence has been considered as part of modified lockdown policy or a gradual lockdown exit strategy. We estimated the impact of such strategies on epidemic and mortality risk using the UK as a case study.

**Methods:** We used an individual based model for a synthetic population similar to the UK, that is stratified into transmission risks from the community, within the household and from other households in the same social bubble. The base case considers a situation where non-essential shops and schools are closed, the secondary household attack rate is 20% and the initial reproduction number is 0.8. We simulate a number of strategies including variations of social bubbles, i.e. the forming of exclusive pairs of households, for particular subsets of households (households including children and single occupancy households), as well as for all households. We test the sensitivity of the results to a range of alternative model assumptions and parameters.

**Results:** Clustering contacts outside the household into exclusive social bubbles is an effective strategy of increasing contacts while limiting some of the associated increase in epidemic risk. In the base case scenario social bubbles reduced cases and fatalities by 17% compared to an unclustered increase of contacts. We find that if all households were to form social bubbles the reproduction number would likely increase to 1.1 and therefore beyond the epidemic threshold of one. However, strategies that allow households with young children or single occupancy households to form social bubbles only increased the reproduction number by less than 10%. The corresponding increase in morbidity and mortality is proportional to the increase in the epidemic risk but is largely focussed in older adults independently of whether these are included in the social bubbles.

**Conclusions:** Social bubbles can be an effective way of extending contacts beyond the household limiting the increase in epidemic risk, if managed appropriately.

## Background

In the UK, similar to many other countries, the introduction of stringent physical distancing measures in March 2020 in response to the Covid-19 pandemic has reduced the transmission of SARS-CoV-19 and alleviated the burden on the healthcare system^1,2^. This reduction, however, has come at great economic, societal, and wider health costs^3–6^. With infection incidence on the decline, countries must now strike a balance between easing restrictions in an attempt to reduce the societal burden while making sure that the epidemic remains under control^7–11^.

Multiple options, that could in combination form an exit strategy, have been proposed to allow easing of restrictions. These include: the widespread use of rapid, potentially app-based, contact tracing in combination with rapid testing and self-isolation^12,13^; expanded random testing to increase detection of asymptomatic infection^14–16^; strict quarantining of travellers on arrival^17,18^; and the use of face masks in high-risk environments^19–22^. Another potential component of a lockdown exit strategy that could allow for greater social interaction is the clustering of contacts beyond the household, commonly referred to as the social bubble or the ‘double bubble’ strategy^23–27^. Under this strategy, households would be allowed to form a cohesive unit with one other household, generating a ‘social bubble’; this would allow individuals to increase their close, physical social interactions beyond their household while potentially limiting the risk of infection through the exclusivity of the bubble. A similar strategy has been implemented in some countries, including New Zealand and Germany, and is currently considered as part of the lockdown exit strategy in the UK^28^.

While physical distancing has placed additional pressures on society as a whole, some households are likely to be disproportionately more at risk of social isolation. Many adults in the UK will have been able to partially shift social contacts online and since 11th May (and 1st June) have been allowed to socialise outdoors with a maximum of one (and subsequently up to five) others while adhering to distancing guidelines^29^. However, such social contact replacements can be more difficult for children, for whom verbal interaction is only a small part of their communication with peers. Further, their carers have often had to balance working from home, childcare and homeschooling, generally without being able to access a support network from family, friends or professional childminders^30^. Single occupancy and single parent households have also likely been disproportionately affected as the complete absence of social face-to-face interactions for many months may impact mental wellbeing^31,32^.

Here, using mathematical models, we assess the likely increase in transmission generated by various plausible social bubble strategies and use the UK as a case study. In particular, we compare the impact of limiting bubbles to those households who would benefit most (single occupancy households and those with young children) with allowing all households to form bubbles. We assess these changes in terms of both the increase in transmission (as characterised by the reproductive ratio, *R*) and short-term increase in fatalities.

## Methods

### Population

The model’s synthetic population was created by generating individuals who are residents of one of 10,000 households. The size of the individual households, as well as the age distribution within households, was sampled to match that observed in the most recent census in England and Wales in 2011 (Figure 1)^33^.

### Transmission model

The transmission dynamics are set to simulate the status of Covid-19 interventions during ‘lockdown’ in the UK in May 2020, in particular simulating contacts that are substantially reduced and largely household-based, with schools, non-essential retail, and leisure facilities closed. This is achieved through stochastic simulation of infections spreading through an interconnected population; connections are captured by a matrix, *A*, which defines the probability that infection can pass between any two individuals in the population. *A* is composed of the sum of two matrices *H* and *B*, which capture within-household and within-bubble transmission respectively (Figure 1). We can then use the matrix *A* to drive forwards the stochastic dynamics using a next generation approach. To this, we add random (mean-field) transmission between individuals in the population to simulate the risk that infection in the wider community poses to the household and the social bubble, and vice versa (Table S1). Transmission rates within the household and the wider community are matched to observed data on secondary attack rates and population-scale R estimates. We assume that households are adhering to current restrictions and social distancing, and therefore largely act as a coherent and largely isolated unit. We therefore assume that the risk of a household acquiring infection from the community is independent of its number of occupants as observed in a cross sectional serological study for SARS-CoV-2 in Germany in March and April 2020^34^.

**Figure 1:**
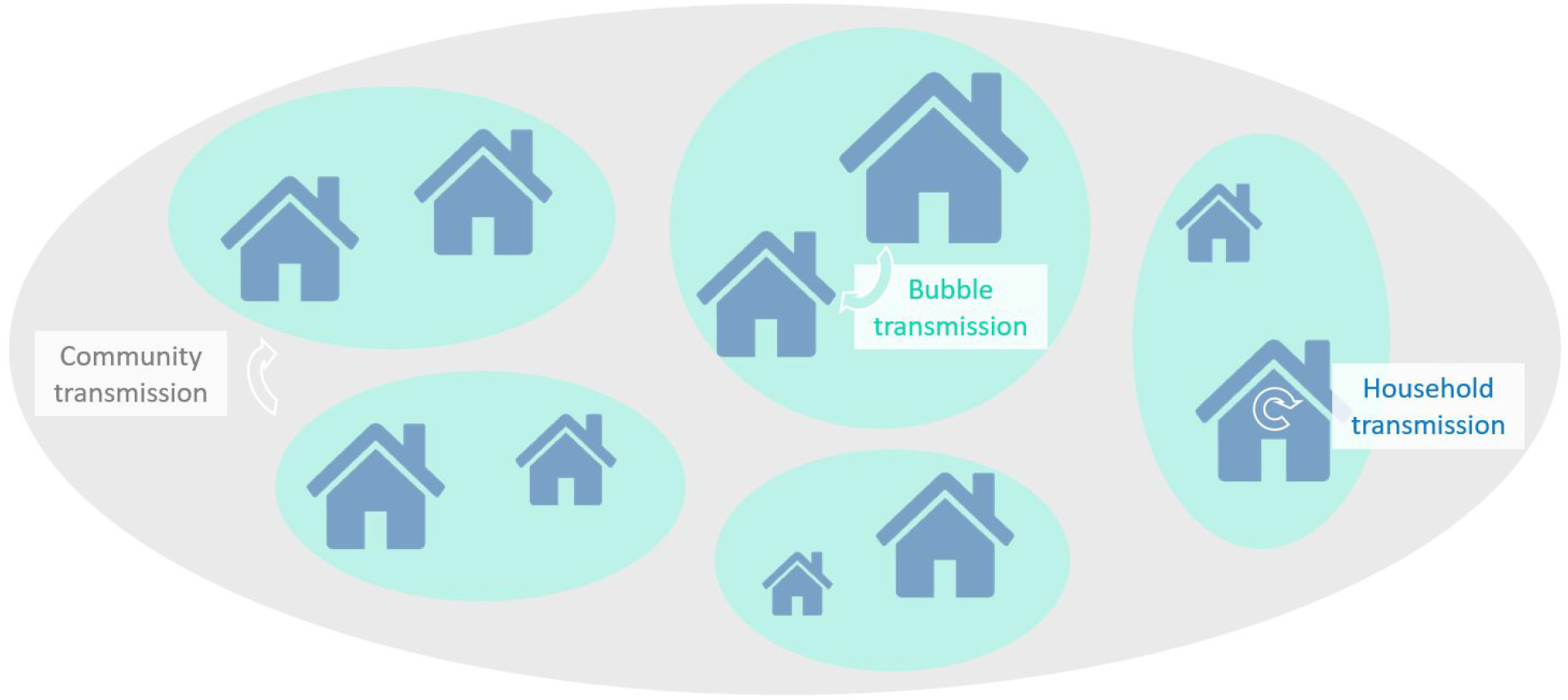
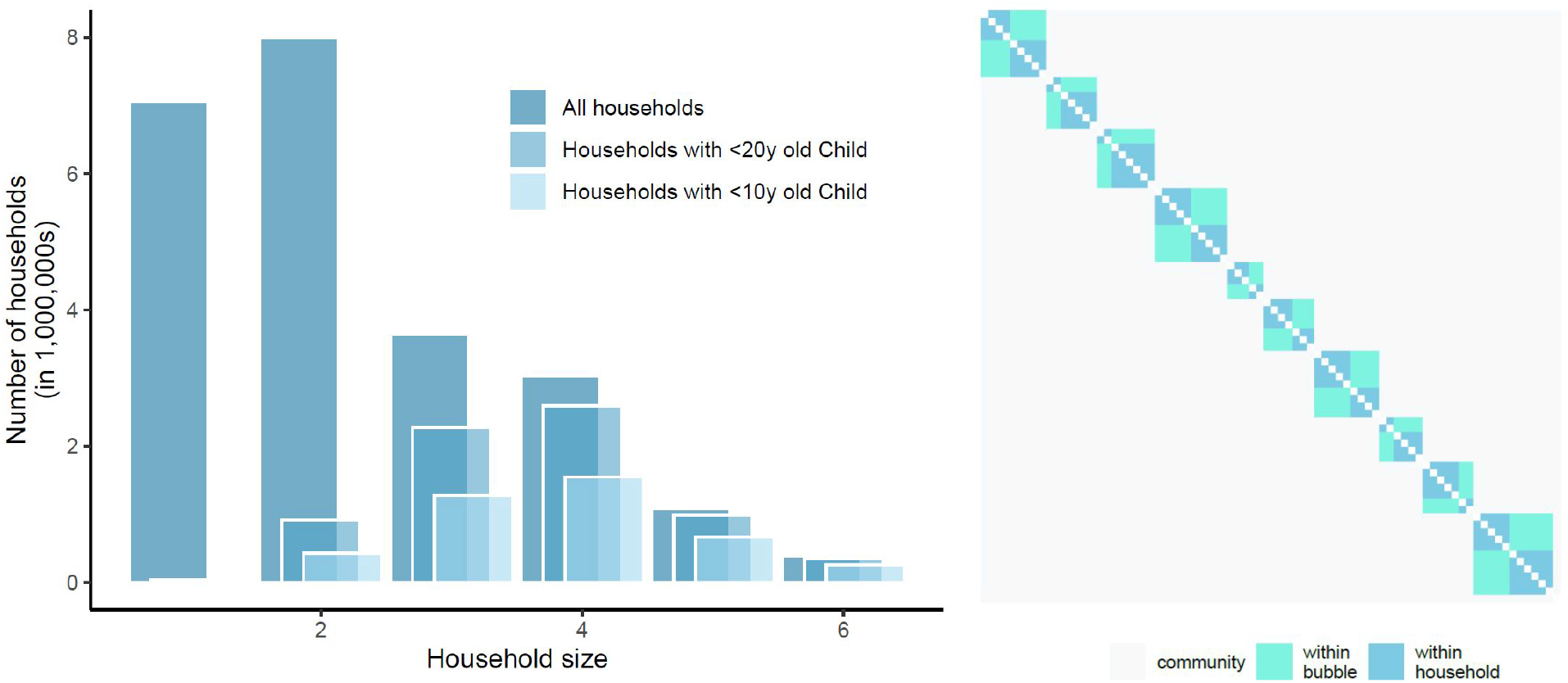
top panel: schematic of model structure and its stratification into different household sizes with three components of transmission dynamics, community transmission, bubble transmission and household transmission; left panel: household size distribution for all households in England and Wales, for those households with at least one child younger than 20 years old and for those with at least one child younger than 10 years old (about primary school age and younger). Right panel: illustrative transmission probability matrix *A*, composed of household and bubble contacts and including community transmission.

We define a baseline household transmission rate, T_H_, as the transmission rate between adults in a two-person household. To this baseline we assume that the susceptibility to infection, *C*, as well as transmissibility of infection, *T*, can be age dependent. There are two conflicting bodies of evidence about the potential role of children. Firstly, it has been observed that children are more likely to experience mild or no symptoms, and as such may have a lower transmission rate^35,36^ (Table S2). Secondly, cases with more severe symptoms are likely to self-isolate reducing their effective infectious period, therefore children that are asymptomatic (or mildly symptomatic) may continue to transmit for longer^34^. In our base parameterisation, we assume that children are 50% as susceptible to infection as adults or elderly adults, but assume that transmissibility is independent of age; this echoes the assumptions of a previous model [33], but an alternative parameterisation based on other work [8] is considered as part of sensitivity analysis. We assume that transmission within households and across households who share a bubble is frequency dependent, i.e. the person-to-person transmission rate within households and across households who share a bubble is inversely proportional to the number of other people in an individual’s household or bubble^34,37,38^.

Throughout, we compare the baseline model without the additional social interactions via bubbles (C1) with different ways in which bubbles could be allowed to form (scenarios 1–6, see below). To assess the effectiveness of social bubbles, compared to increasing contacts in an unclustered fashion, we consider two further comparison scenarios. In C2, individuals form the same number of additional contacts with the population as in Scenario 6, but these are chosen randomly across the population. In C3, the community transmission rate is increased to match the total increase in the transmission rate in Scenario 6. Scenario 6, C2, and C3 therefore represent fixed and clustered, fixed and unclustered, and variable additional contacts respectively.

For a technical summary of the model and its assumptions, see the supplemental material.

### Outcome metrics

We calculate two key metrics for the epidemiological impact of interventions in our household model with extended social contacts, which relate to epidemic risk and adverse health measures.

The net reproduction number (*R*) is a measure of risk for (increased) transmission that may eventually result in an exponential increase in infections and hence the need for stricter control measures if exceeding the epidemic threshold (*R>1*). *R* is defined as the number of secondary infections generated by a typical case. In models incorporating household structure, the typical case is effectively an average over the probability that such a case is the first, second, third or later generation case within the household. Following the principle of Pellis et al.^39,40^, we determine *R* numerically as the ratio of the number of new infections in the fifth to the fourth model generation, adjusted to account for the partial depletion of susceptibles. In all simulations this provided sufficient time for the average state of infectious individuals to have stabilised without a notable depletion of susceptibles in the overall population (Supplementary Figure S1).

Our second metric is the relative mortality (i.e. number of deaths), compared to the baseline model (C1) of isolated households; this provides a measure of adverse health impacts as a result of increased contact rates in the respective scenarios. We use age stratified infection fatality rates (IFR) estimated from repatriation flights early in the Covid-19 pandemic^41,42^ to predict the mortality risk in the five generations following model burn in (i.e. from the fourth to ninth model generation – approximately the second month after social bubbles were initiated).

### Parameterisation

To parameterise the Covid-19 transmission dynamics in the model we need to define the infection dynamics within a household, within a bubble and from the community. To parameterise the within household transmission we assume that, in line with observations from contact tracing while accounting for some underreporting^43–45^, the secondary household attack rate (SAR_HH_) is 20%. This is achieved by tuning the transmission rate (*τ_H_*) between household members to achieve this average attack rate. We subsequently assume that community transmission is such that, in combination with household transmission, the model generates an overall reproduction number of 0.8, similar to estimates from mid-May 2020 in the UK^46,47^. Further, as a base-case, we assume that transmission between households within the same bubble is 50% lower than that within a household, i.e. *T_B_* = ½* *T_H_*. In our base case parameterisation a 3.2-fold increase in community contacts yielded a reproduction number of about 2.5; this is in line with an approximate 70% reduction in contacts during lockdown and a reproduction number of about 2.5 in the early phase of the pandemic with hardly any distancing measures in place^46^.

We additionally assume that all eligible households would take up the opportunity to expand their contacts and enter into a social bubble with one other household, and that they would adhere to the exclusivity of this bubble. The impact of only partial uptake is explored in the Supplementary Figure S2, where we find that the increase in *R* scales approximately linearly with uptake. The impact of adherence, incorporated by allowing 50% of eligible households to form an additional social bubble, is explored in our sensitivity analyses (Figure 4; Supplementary Figure S3-S7).

All analyses were done in Matlab^48^ and R^49^ and are available via github on github.com/tsleng93/SocialBubble

### Scenarios Modelled

We considered a number of contact clustering strategies of how bubbles could be allowed under any relaxation to lockdown measures:

1. Allow households with children younger than 10 years old (about primary school age or younger) to pair up
2. Allow households with children younger than 20 years old to pair up
3. Allow single occupancy households to pair up with another single occupancy household
4. Allow single occupancy households to pair up with another household of any size
5. A combination of scenarios 1 & 3
6. Allow all households to pair up with one other household

All these scenarios assume that the pairing will occur at random between permitted households.

We compare the above scenarios against three counterfactuals that do not include social bubbles. These allows us to elucidate the impact of

C1) Perfect adherence to the current household-only contact strategy (other than the background transmission risk from the community)
C2) All individuals increase their number of contacts so that population level force of infection matches that of Scenario 6. Contacts are unclustered and chosen at random across the population but stay the same over time.
C3) All individuals increase their number of contacts so that population level force of infection matches that of Scenario 6. Contacts are unclustered and chosen at random across the population and are re-sampled at each generation.

Counterfactuals 2 and 3, maintain the same level of additional contacts outside the home as social bubbles but change how they are distributed.

### Sensitivity analyses

Other than the previously described base case we performed a number of univariate sensitivity analyses to test the robustness of our findings to the underlying assumptions. Specifically, we assume that the current *R* is 0.7 or 0.9 instead of 0.8^47^; that the secondary attack rate in the household is 10% or 40% instead of 20%^44^; that transmission between individuals in the same bubble (but different households) is 10% or 100% of that within a household instead of 50%; that the risk of a household to get infected with SARS-CoV-2 from the community increases with increasing household size instead of being independent; that 50% of bubbles do not adhere to the recommendations but also form bubbles with an additional household rather than perfect adherence; and that the relative susceptibility to infection of children and older adults compared to adults is 79% and 125% while the relative transmissibility is 64% and 290%, respectively^8,50^.

## Results

### Households

From the 2011 census of England and Wales, the average size of a household was 2.32 persons. Considering households with at least one child under 10, the average household size increases to 3.77 persons, and 29.3% of the population live in such households. For households with at least one child under 20, the average household size is 3.65 persons, and 48.5% live in such households. In total, only 15% of households are occupied by someone over the age of 60, although 50% of single occupancy households were occupied by such older adults. There is limited multi-generational mixing, with only 3% of households having both a child under 10 and an adult over 60.

### Impact of social bubble strategies on epidemic risk

Assuming an initial reproduction number of 0.8, perfect adherence to the recommended social bubble strategy and that all eligible households indeed pair up, we find that strategies that exclusively target single person households (scenario 3) or households with young children (scenario 1) do not increase transmission substantially (*R* of 0.84 and 0.86 respectively in the base case scenario); their combination (scenario 5) is also predicted to only marginally increase transmission in the community (*R* of 0.90) (Figure 2). For these two targeted strategies, even under conservative assumptions (SAR_HH_ = 40%, T_H_ = T_B_), the increase in transmission is unlikely to lead to substantial spread of Covid-19 (*R* of 0.89 and 1.01 for scenario 1 and 3, respectively). However, allowing all households to form bubbles (scenario 6) is estimated to increase the reproduction number to 1.09 and hence substantially beyond the critical threshold value of 1 for the base case scenario (Figure 2).

**Figure 2:**
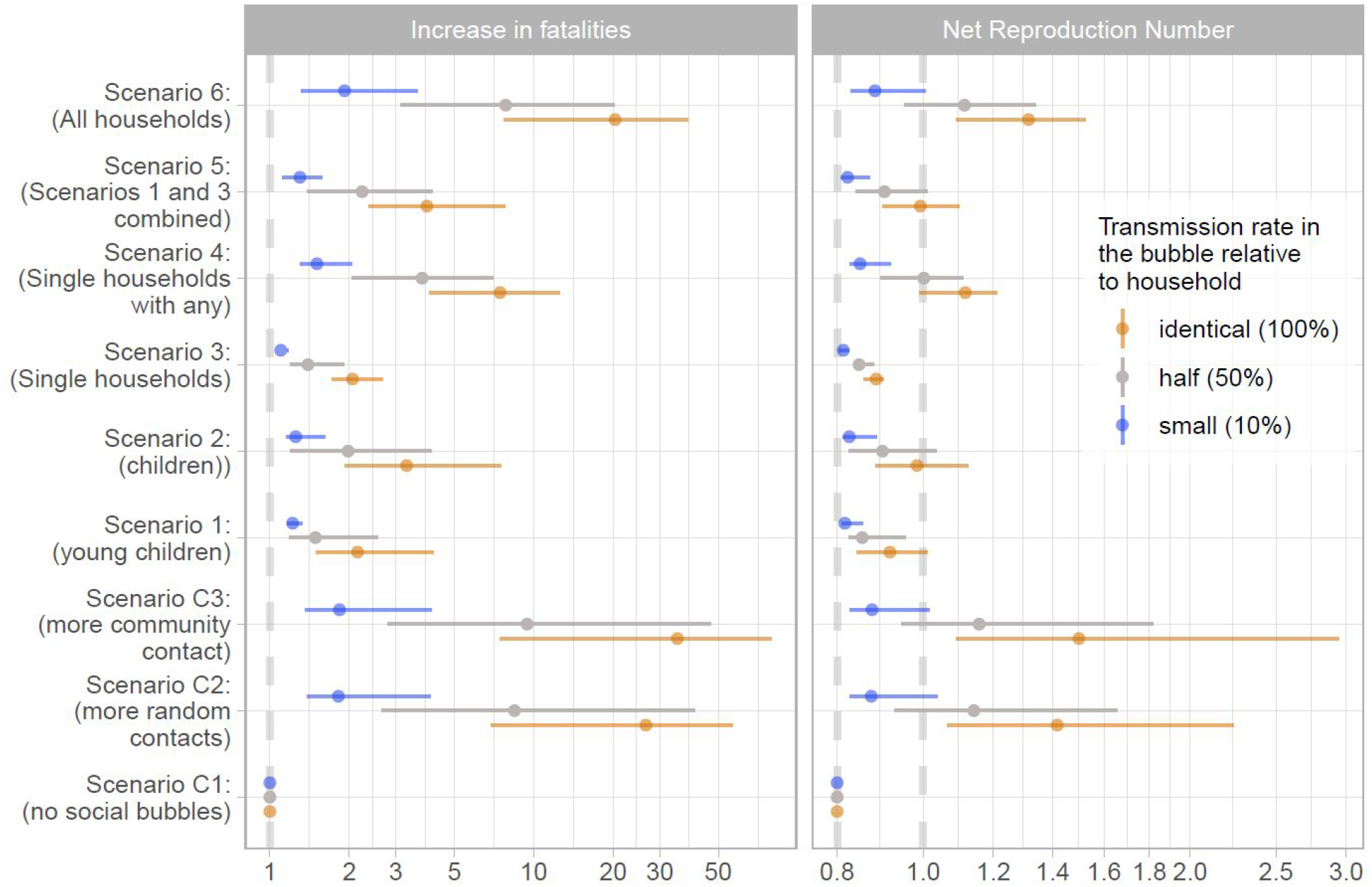
Estimated reproduction number and increase in fatalities for the considered scenarios under the assumption that all eligible households pair up and thereby form exclusive social bubbles and that transmission rates within a social bubble are the same as within the household. Central estimates are assuming SAR_HH_ = 20% and the upper and lower limits represent the respective 10% and 40% assumption.

Generally, the fewer households that were deemed eligible for expanding their social bubble under a specific strategy, the smaller the average household size of those involved, the smaller the uptake (see Supplementary Figure S2) and the smaller the risk for transmission within the bubble, the smaller the increase in transmission as a result.

### The impact of social bubble strategies on mortality risk

The average age in the households eligible to form social bubbles in scenarios 1 to 6 was 22.2, 25.8, 58.7, 41.0, 33.5, and 40.2 years, hence the average infection fatality risk in an average household member implementing such a strategy was 0.10%, 0.14%, 2.49%, 1.08%, 0.84%, and 0.97%. In all scenarios, the increased number of contacts lead to both excess infections and fatalities. Excess risk for infection compared with no social bubbles (Scenario C1) was seen in households implementing the social bubbles as well as those households who were not eligible, although, as expected, the relative risk for infection was higher in eligible households (Figure 3). The resulting excess mortality risk depended highly on the estimated epidemic risk but also on the average age of the affected households. For example, social bubbles among households with young children (Scenario 1) and among single occupancy households (Scenario 3) led to similar increases in infections, risk ratio of 1.64 and 1.50, respectively, in the base case scenario. While the resulting overall increase in mortality risk was similar in both scenarios (Figure 2) the mortality risk in Scenario 1 was largely attributed to households not eligible to form social bubbles (Figure 3)

**Figure 3:**
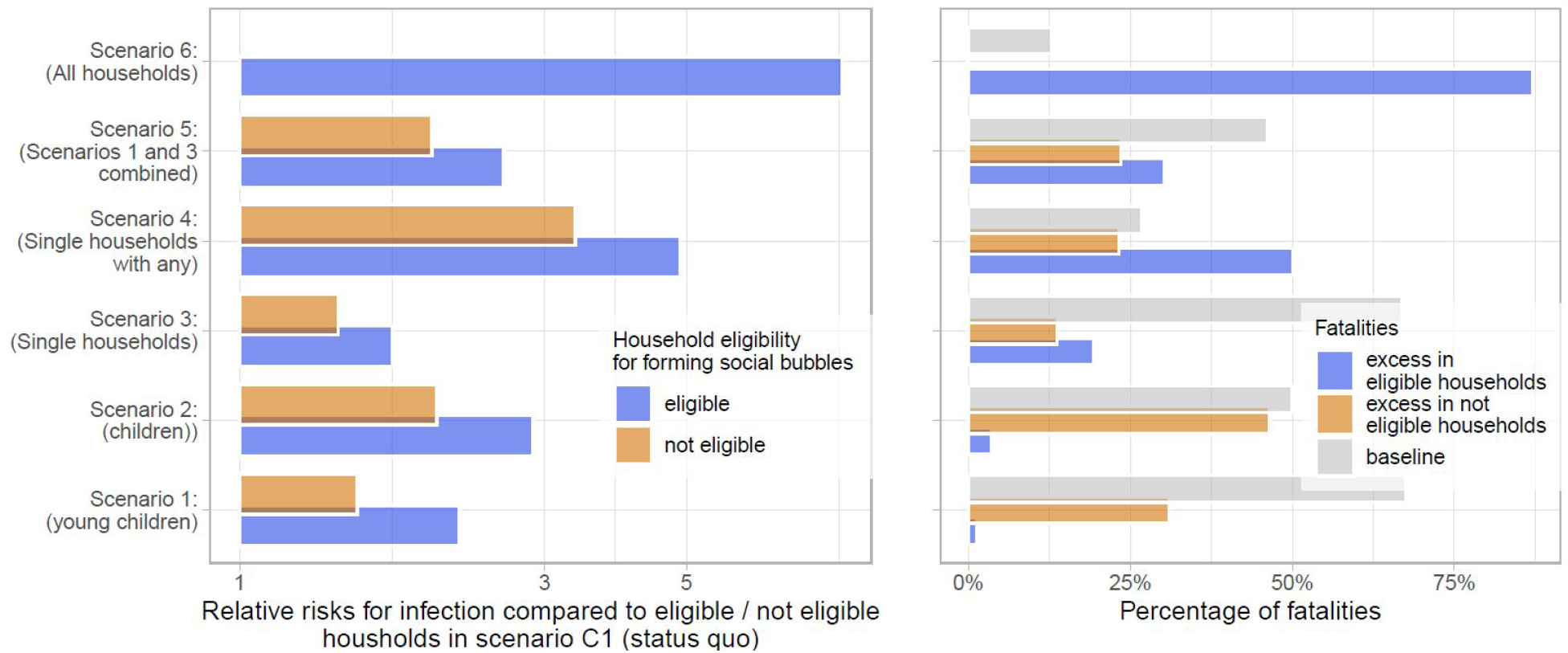
Relative risk of infection and fatality. Left panel: the relative risks for infection in the considered scenarios if compared to the status quo with no social bubbles (Scenario C1), stratified into the risks in households eligible and not eligible for forming social bubbles. Right panel: the population attributable fraction of fatalities in the considered scenarios. The overall mortality risk is stratified into the baseline risk, and the excess risk from forming social bubbles in both eligible and non eligible households.

The forming of social bubbles was effective at reducing the infection and thereby the mortality risk compared to strategies that increased contacts in a less clustered way: under base case assumptions all households forming social bubbles (Scenario 6) reduced the mortality risk by 7.5% and 17.1% compared to adding the same amount of contacts randomly (Scenario C2) and time varying (Scenario C3).

### Sensitivity analyses

We tested the robustness of our findings to a number of alternative assumptions governing the spread of SARS-CoV-2 and the implementation of the social bubble strategy. Within the tested parameter space, the alternative assumptions did not qualitatively change our findings. The two main factors that increased or decreased the epidemic risk were an initial value of *R* closer to one when implementing the strategy and a much higher than typically observed secondary household attack rate. However for strategies 1 and 3, in neither of the univariately tested scenarios did *R* exceed one (Figure 4 & Supplementary Figure S3). The assumptions on age stratified susceptibility and transmissibility were conservative for strategies focussed on households with children and were optimistic for single-person households; and vice versa for the assumption that the risk for community transmission was independent of household size (Supplementary Figures S3-S7). The epidemic risk from social bubbles is further reduced if within bubble transmission is reduced to 10% of that within household transmission.

**Figure 4:**
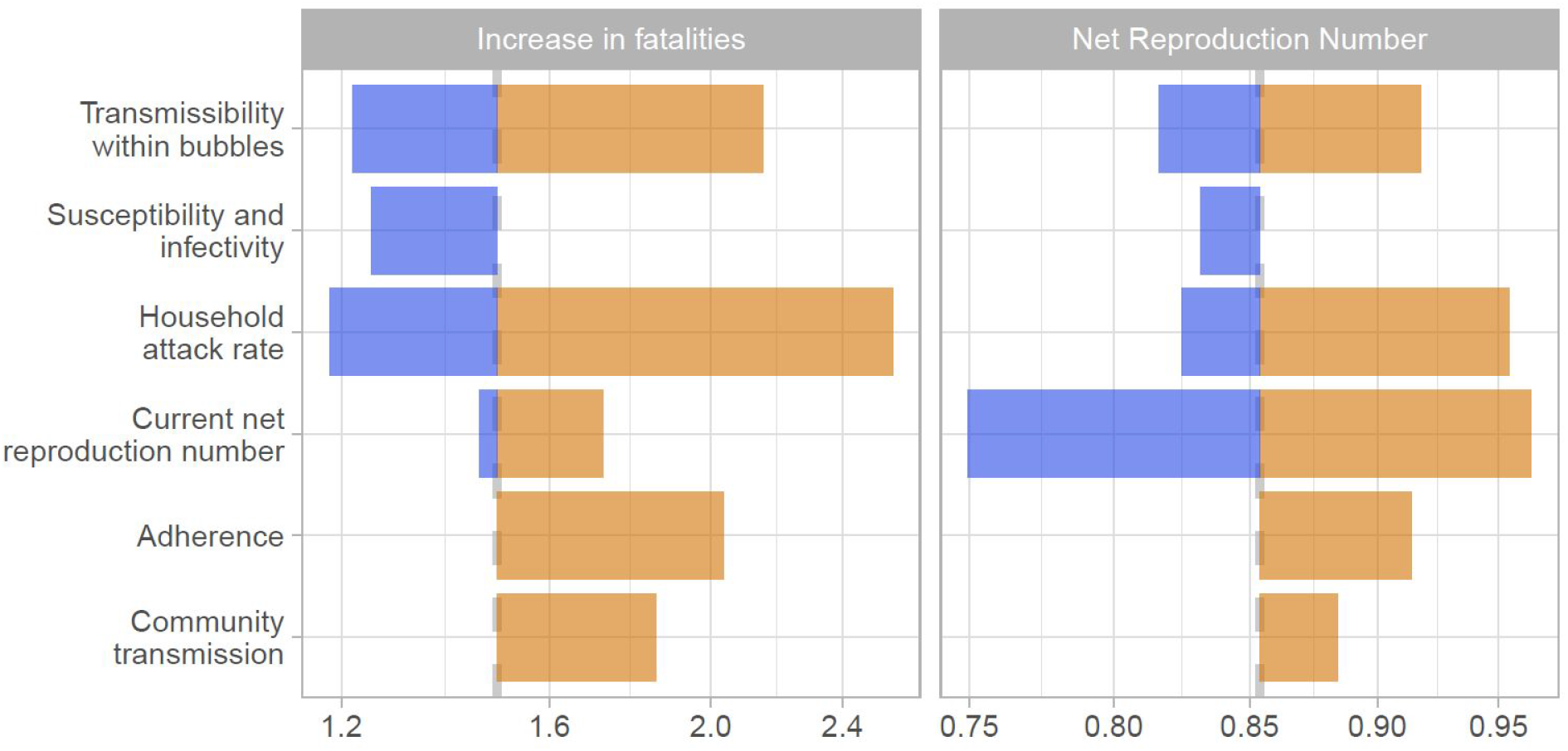
Sensitivity analyses. The tornado diagram shows a univariate sensitivity analysis on the expected increase in fatalities and the net reproduction number for scenario 1, i.e. allowing households with young children to pair up. Increases over the base case estimate are shown in orange and decreases in blue. The base case estimate is indicated through the dashed grey vertical line. The sensitivity scenarios are (from top to bottom): transmission across individuals of households sharing a bubble is 90% or 0% lower than that within a household instead of 50%; the relative susceptibility to infection of children and older adults compared to adults is 79% and 125% while the relative transmissibility is 64% and 290%; the secondary attack rate in the household is 10% or 40% instead of 20%; *R_e_* is 0.7 or 0.9 instead of 0.8; that 50% of bubbles not adhere to the recommendations but pair up to form bubbles with four households, rather than perfect adherence; and that the risk of a household to get infected from the community is proportional to the household size instead of being the same across households.

The effectiveness of social bubbles also varied according to the Scenario considered and the parametric assumptions and was as large as a 27.2% and a 70.0% reduction in mortality risk compared to adding the same amount of contacts randomly (Scenario C2) and time varying (Scenario C3), under base case parameters except for the age dependent susceptibility and infectivity of cases.

## Discussion

We found that contact clustering, or the forming of social bubbles that join two households, can allow increased social contacts beyond the households while limiting additional risk for transmission. In the base case social bubbles reduced the mortality risk by 17% compared to a scenario that increased contacts by the same amount but without clustering thereof, and risk reduction under some alternative parameterisations was even higher. Allowing all households to form social bubbles may increase the reproduction number above its epidemic threshold and hence lead to an increase in cases. A strategy that sees only those at potentially the highest need for an extension of their contacts beyond the household (families with young children and single-person households) should lead to a limited increase in epidemic risk (less than 10% individually and less than 15% in combination) which remained below the epidemic threshold in most scenarios considered. The epidemic risk can be further reduced if the transmission risk within the bubble is minimised. As the number of contacts and *R* increase with a social bubble strategy, so does the risk of adverse health outcomes. We find that adverse health outcomes are largely proportional to the epidemic risk, but will disproportionately affect households with older adults independently of their clustering behaviour.

Stringent physical distancing policies in many countries have reduced the reproduction number from about 2.5 to just under 1^11,46,51^. This provides the opportunity to risk a small amount of additional contacts without necessarily experiencing an increase in Covid-19 cases, if crossing the epidemic threshold can be avoided. Here, we investigate the effectiveness of social bubbles as a potential option to ease the social impact of the lockdown without increasing transmission risks. However, while we here look at the impact of social bubbles in isolation these would only be part of a multi-variable exit strategy^28^. Hence, our comparisons of alternative bubble strategies against the epidemic threshold should be interpreted cautiously and in consideration of the other changes to behaviour, such as re-opening of non-essential retail and travel. It is likely that these other activities will combine in a non-linear manner so that bubbling in a context of children returning to school might affect results.

Countries including Germany and New Zealand have implemented strategies similar to those considered here. In Bavaria, Germany, in early May and before the reopening of schools and nurseries, up to three households could form exclusive groups to share childcare amongst them^52^. Even during their highest national alert level, level 4 “Lockdown”, New Zealand permitted people living alone to pair up with a “lockdown buddy” and key workers to identify “childcare buddies”. New Zealand moved to level 3 in their Covid-19 alert system, “Restrict”, on 27 April 2020 which included the advice to residents to stay within their household bubbles but permitted expansion of such to reconnect with close family, bring in caregivers or support isolated people^53^. A subsequent survey found that among respondents the highest increase in the quality of life by far would not be brought by re-opening of schools, shops, churches or fitness centres, but by allowing households to re-connect^54^. It also found that in going to alert level 3 only 50% of households took up the opportunity to expand their social bubbles and that there was high awareness of the importance of the exclusivity of the bubbles, with only 7.5% of bubbles reporting to have had contacts outside their bubble.

We identify three key risks to the success of social bubbles that may increase their epidemiological risk: potential lack of adherence, a higher than observed secondary household attack rate and being too close to the epidemic threshold. If the risk perception of the population changes as a result of allowing parts of the population to form social clusters, a lack of adherence to the exclusivity of the bubbles could lead to rebuilding of contact networks that in turn lead to the epidemic threshold being crossed. We find that some degree of non-adherence would not necessarily hinder the success of the strategy, but communication of the strategy is likely to be key. For example, in New Zealand, the social bubbles were not framed as a relaxation of social distancing rules but rather as a source of support for those who are at a higher risk of social isolation or with needs for care, including childcare^54^. We find that if the secondary household attack rate is substantially higher than assumed in our base case the epidemic risk is elevated close to the epidemic threshold. While high household attack rates have been observed in some instances, our base case assumptions are in line with an increasingly consistent picture emerging in the academic literature^44^. Also, superspreading events have been raised as a potentially important source for sustained transmission of SARS-CoV-2, which would further imply a rather low secondary household attack rate in most instances^55,56^. Similarly, if the reproduction number is very close to its epidemic threshold, an increase in contacts, even if clustered, could result in an increase in cases. Hence, careful monitoring of such is needed to assess the feasibility of expanding social bubbles.

An expansion of contacts into social bubbles will naturally lead to some increase in transmission in comparison to perfect adherence to the recommendation to restrict to all but essential contacts outside the household. However, such adherence may decline as a result of extended periods of time in lockdown and lead to an expansion of contacts that are unclustered, potentially leading to long chains of transmission. To illustrate such a scenario, we include alternative comparisons for the strategy that allows all households to form social bubbles (Scenario 6). We considered strategies that would have the same overall increase in transmission as in that scenario but where either the contacts or not cluster but stay fixed over time (Scenario C2) or where contacts are not clustered and vary over time (Scenario C3). We show that the clustering slightly reduces the epidemic and reduces the number of infections and subsequent fatalities by 5% and 17% in the base case and even more in some of the parametric sensitivity analysis. Hence social bubbles, if given as a guidance to households who are struggling to cope with the lockdown, may give these households a safer alternative and thereby help to reduce the epidemic and mortality risk. This may particularly be the case for households with single parents or parents who cannot easily work from home; in such circumstances allowing social bubbles may help increase equity in the impact of the lockdown.

Our analyses have a number of limitations. Firstly, we only assessed the risk of extending social bubbles but not the benefits. As of June 2020 in England, social contact beyond the immediate household is restricted to virtual contact or contact in open spaces with up to 5 individuals while keeping 2 meters apart. In other words, one can have a conversation. While conversations are a large part of the social contacts of adults they have little role in the social interactions of young children. Hence the benefit of extending bubbles for children is likely disproportionately higher. Furthermore, clustering contacts into social bubbles is likely to ease contact tracing which is an integral part of both containment and lockdown exit strategies. We considered social bubbles against the background of a lockdown, particularly where schools are closed. As lockdown measures are eased and schools are gradually re-opened forming social bubbles that largely overlap with societal one (for example forming social clusters with families that have children going to the same class) is likely further reducing the additional epidemic risk from social bubbles. We also did not include the possibility to form bigger social bubbles that would cluster together 3 or more households. While this has been implemented in other countries, the complexity of creating an exclusive cluster of three or more households could lead to a loss of adherence. We did not consider further heterogeneity of the society. This may include that about 20% of the working population is classified as key workers and will have an increased risk for infection from the community, or that families with lower socioeconomic status have been hit disproportionally hard by the lockdown. Further, we also did not consider age-homogeneous mixing when pairing up single occupancy households and subsets of such strategies, that may exclude high risk individuals and thereby reduce the mortality risks in that strategy.

## Conclusions

Our analyses highlight the continued need for social distancing despite a social bubble strategy being an effective way to expand contacts while limiting the risk for a resurgence of cases. Recommending social bubbles only for those who particularly struggle with the lockdown, while minimising opportunities for spread through prioritising outdoor settings for gathers and adhering to distancing recommendations as possible, may strike an effective balance between minimising the impact on mental health and minimising the epidemic Covid-19 risk.

## Data Availability

The simulation model and analyses from this study are available via github on  github.com/tsleng93/SocialBubble

## Acknowledgements

The following authors were part of the Centre for Mathematical Modelling of Infectious Disease 2019-nCoV working group. Each contributed in processing, cleaning and interpretation of data, interpreted findings, contributed to the manuscript, and approved the work for publication:

Sophie R Meakin, Gwenan M Knight, Kiesha Prem, Damien C Tully, Graham F. Medley, Billy J Quilty, Eleanor M Rees, Sebastian Funk, C Julian Villabona-Arenas, Kevin van Zandvoort, Jon C Emery, Amy Gimma, David Simons, Anna M Foss, Petra Klepac, W John Edmunds, Mark Jit, James D Munday, Stéphane Hué, Joel Hellewell, Hamish P Gibbs, Samuel Clifford, Sam Abbott, Christopher I Jarvis, Katherine E. Atkins, Matthew Quaife, Akira Endo, Georgia R Gore-Langton, Charlie Diamond, Timothy W Russell, Megan Auzenbergs, Alicia Rosello, Carl A B Pearson, Kathleen O’Reilly, Fiona Yueqian Sun, Nikos I Bosse, Quentin J Leclerc, Rein M G J Houben, Thibaut Jombart, Simon R Procter, Arminder K Deol, Emily S Nightingale, Rosalind M Eggo, Rachel Lowe, Yang Liu.

The following funding sources are acknowledged as providing funding for the working group authors. Alan Turing Institute (AE). BBSRC LIDP (BB/M009513/1: DS). This research was partly funded by the Bill & Melinda Gates Foundation (INV-003174: KP, MJ, YL; NTD Modelling Consortium OPP1184344: CABP, GFM; OPP1180644: SRP; OPP1183986: ESN; OPP1191821: KO’R, MA). DFID/Wellcome Trust (Epidemic Preparedness Coronavirus research programme 221303/Z/20/Z: CABP, KvZ). Elrha R2HC/UK DFID/Wellcome Trust/This research was partly funded by the National Institute for Health Research (NIHR) using UK aid from the UK Government to support global health research. The views expressed in this publication are those of the author(s) and not necessarily those of the NIHR or the UK Department of Health and Social Care (KvZ). ERC Starting Grant (#757688: CJVA, KEA; #757699: JCE, MQ, RMGJH). This project has received funding from the European Union’s Horizon 2020 research and innovation programme – project EpiPose (101003688: KP, MJ, PK, WJE, YL). This research was partly funded by the Global Challenges Research Fund (GCRF) project ‘RECAP’ managed through RCUK and ESRC (ES/P010873/1: AG, CIJ, TJ). HDR UK (MR/S003975/1: RME). Nakajima Foundation (AE). NIHR (16/137/109: BJQ, CD, FYS, MJ, YL; Health Protection Research Unit for Modelling Methodology HPRU-2012-10096: TJ; PR-OD-1017-20002: AR). Royal Society (Dorothy Hodgkin Fellowship: RL; RP\EA\180004: PK). UK DHSC/UK Aid/NIHR (ITCRZ 03010: HPG). UK MRC (LID DTP MR/N013638/1: EMR, GRGL, QJL; MC_PC 19065: RME; MR/P014658/1: GMK). Authors of this research receive funding from UK Public Health Rapid Support Team funded by the United Kingdom Department of Health and Social Care (TJ). Wellcome Trust (206250/Z/17/Z: TWR; 208812/Z/17/Z: SC; 210758/Z/18/Z: JDM, JH, NIB, SA, SFunk, SRM). No funding (AKD, AMF, DCT, SH).

## Supplement to The effectiveness of social bubbles as part of a Covid-19 lockdown exit strategy, a modelling study

Trystan Leng, Connor White, Joe Hilton, Adam Kucharski, Lorenzo Pellis, Helena Stage, Nick Davies, CMMID-Covid-19 WG, Matt Keeling, Stefan Flasche

### Further details on the methods

In this study we use a stochastic network simulation model to assess the impact of social bubbles. Here, we describe the model in detail. Table S1 describes our notation, while Table S2 contains the formulae for transmission rates within our model.

**Table S1:** Model notation

| Symbol | Meaning |
| --- | --- |
| $a_i$ | Age group of an individual $i$, $a_i \in \{\text{Child, Adult, Older adult}\}$ |
| $C(a)$ | Age-dependent susceptibility scaling factor |
| $T(a)$ | Age-dependent transmissibility scaling factor |
| $N_H(i)$ | Number of individuals in an individual $i$'s household |
| $N_B(i)$ | Number of additional individuals in an individual $i$'s bubble outside of their household, i.e. the size of household joined to |
| $\tau_H$ | Baseline transmission rate across household contact |
| $\rho_H(i,j)$ | Household transmission rate from individual $j$ to individual $i$ |
| $\tau_B$ | Baseline transmission rate across bubble contact, $\tau_B = c\tau_H$ where $c \in [0, 1]$ |
| $\rho_B(i,j)$ | Bubble transmission rate from individual $j$ to individual $i$ |
| $\varepsilon$ | Baseline mean-field transmission rate |
| $\varepsilon(i)$ | Mean-field transmission rate to an individual $i$ |
| $\mathbf{l}(g)$ | Vector of infection statuses of individuals at generation $g$ (including those who have now recovered) |
| $I(g)$ [$I_a(g)$] | Number of infected individuals [of age $a$] at generation $g$. |
| $S(g)$ | Number of susceptible individuals at generation $g$. |
| $N$ | Size of population. |

**Table S2:** Transmission rates

| Transmission rate | Model formulation |
| --- | --- |
| Household, $\rho_H(i,j)$ | $\frac{T(a_j)C(a_i)\tau_H}{N_H(i)-1}$ if $i$ and $j$ are within the same household, 0 otherwise |
| Bubble, $\rho_B(i,j)$ | $\frac{T(a_j)C(a_i)\tau_B}{N_B(i)}$ if $i$ and $j$ are within the same bubble (but not the same household), 0 otherwise |
| Mean-field, $\varepsilon(i)$ | $\frac{C(a_i)\varepsilon}{N_H(i)N} \sum_a T(a)(I_a(g) - I_a(g-1))$ |

The baseline household transmission rate, *τ_H_*, is defined as the transmission rate between adults in a household of size two. Similarly, the baseline bubble transmission rate, *τ_B_*, is defined as the transmission rate between households in a bubble consisting of two adults. Specific transmission rates across household (or bubble) contacts are inversely proportional to the number of other people in an individual’s household (to the number of people in an individual’s bubble). They also depend upon the transmissibility of the individual *j* transmitting infection (T(a_j_)), and upon the susceptibility of the individual *i* receiving infection (C(_i_)), which are dependent on the age classes of individuals *j* and *i*. The mean-field transmission to an individual is inversely proportional to the number of individuals in their household, as we assume that a household acts as a coherent and largely self-contained unit when interacting with the population at large. *ε(i)* also depends on the susceptibility of *i*, determined by their age class. The force of infection from the general population is given by ΣT(a)(I_a_(g) – I_a_(g-1)), i.e. the new infections in generation g of each age class a, scaled by the relative transmissibility of that age class.

By considering transmission as a Poisson process, we obtain the elements of the probability matrices H and B, the matrices of within household and within bubble transmissions respectively, by taking H(i,j) = 1-e^−⍴H(i,j)^and B(i,j) = 1-e^−⍴B(i,j)^. A non-zero element within the matrix H (or B) indicates that the corresponding individuals are within the same household (or bubble). We obtain the overall probability matrix for the population by taking A = H+B.

In order to simulate an epidemic, we begin by randomly sampling the probability matrix A. Doing so, we retain only the infectious connections between individuals that will lead to an infection. We refer to the sampled matrix as A’. A’(i,j) = 1 denotes that individual *j* will infect individual *i* with probability 1, given individual *j* is infected. We initiate each simulation with 100 infectious individuals chosen uniformly at random from the population. Letting **<u>I(g)</u>** be the vector of infection statuses of individuals in generation *g*, we obtain the next generation by **<u>I(g+1)</u>** = sign((A’+Id)’\***I(g)**), where Id is the identity matrix, and where sign() is an element-wise function equal to 1 for each positive element and 0 otherwise. Via this matrix multiplication, every newly infected individual in generation *g* infects all of their infectious contacts that generation. Here the identity matrix is added to impose that individuals do not become susceptible again after one generation, while the sign function is used to impose that individuals cannot be infected more than once. This process can be iterated until equilibrium is reached, and the epidemic has ended. To this, we also add mean-field transmission. Each generation, the number of new infections is calculated in order to calculate *ε(i)* for each susceptible individual i, who is infected from mean field transmission with probability 1 – e^ε(i)^ each generation.

Recovery from infection is not explicitly modelled in the simulation, but rather is implicitly built into the structure of the model. If an individual *i* is infected in generation g, they will infect all of their transmission contacts in generation *g* +1 via the matrix multiplication. They also only contribute to community infection in generation *g* +1. While individual *i* remains ‘infected’ (with value 1), they no longer play any role in the infection dynamics, nor can they be reinfected. Hence, the simulation model assumes that individuals are infectious for one generation, before recovering with immunity.

Results are averages obtained from simulations of 100 epidemics for 10 different sampled epidemic networks, hence results are averages of 1000 simulations.

While in this study, we consider households and the effect of introducing bubbles to an epidemic, and consider three age classes (children, adults, and older adults), our simulation methods are general, and could be used for an arbitrary probability matrix with an arbitrary number of risk classes. The simulation model and analyses from this study are available via github on github.com/tsleng93/SocialBubble.

The social bubble strategy works under the assumption that bubbles are exclusive. We model non-adherence to the strategy by allowing 50% of eligible households to enter into close contact with an additional household. Doing so means that bubbles are no longer mutually exclusive, and that chains of transmission could potentially span many households. Letting B_2_ denote the probability matrix of additional bubbles through non-adherence, A is now obtained by the sum of H, B, and B_2_.

As well as comparing our results to the baseline scenario where households do not form bubbles, we also consider two counterfactual situations, C2 and C3, where individuals have increased their social contacts by an equivalent amount to Scenario 6 (where all households have entered into social bubbles). In C2, we consider a situation where each individual has the same number of fixed additional contacts as in Scenario 6, but contacts are chosen at random across the population instead of being concentrated in one another household. The rate matrix B for C2 is obtained by taking the rate matrix B for Scenario 6, and swapping links with probability 1.

In C3, we consider a situation where each individual has increased their number of contacts, and that these contacts are not necessarily fixed. This is achieved by increasing each individual’s rate of mean-field transmission to match the additional rate of transmission included from bubble contacts in scenario 6. This new rate of mean-field transmission, ε*, is given by finding the value satisfying:

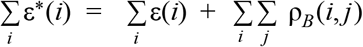

Rearranging, we can obtain an expression for ε*:

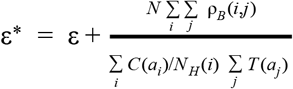

Hence, the total rate of additional transmission to an individual from outside of their immediate household is matched between C3 and Scenario 6.

### Outcome metrics

The primary outcome metric we consider in this paper is the net reproduction number, R. While the standard method of calculating *R* for is to take the dominant eigenvalue of the next generation matrix^57^, such a method fails when considering a population with households. This dominant eigenvalue does not possess the threshold qualities *R* implies, and does not account for the early local depletion of susceptibles household structure imposes. Accordingly, a variety of metrics have been proposed^58^. However, even in simpler models than ours, the formal derivation of *R* for households can be involved. Therefore, we take a numerical approach to calculating *R*. We take the ratio between new infected individuals in the fifth and fourth generations, adjusted for the depletion of susceptibles. Specifically, defining *R(g)* as *R* = *(I(g+1)/I(g)) × (N/S(g)*), we take *R(4)* as *R*. We observe that this function *R* reaches an equilibrium after this many generations, which persists for a number of generations. This is demonstrated in Supplementary Figure S1, and for analyses we check that *R* has stabilised by considering whether *R(5)* is within 5% of *R(4)*. If *R* has not stabilised, we rerun the simulation with 10, instead of 100, initially infected individuals.

**Supplementary Figure S1:**
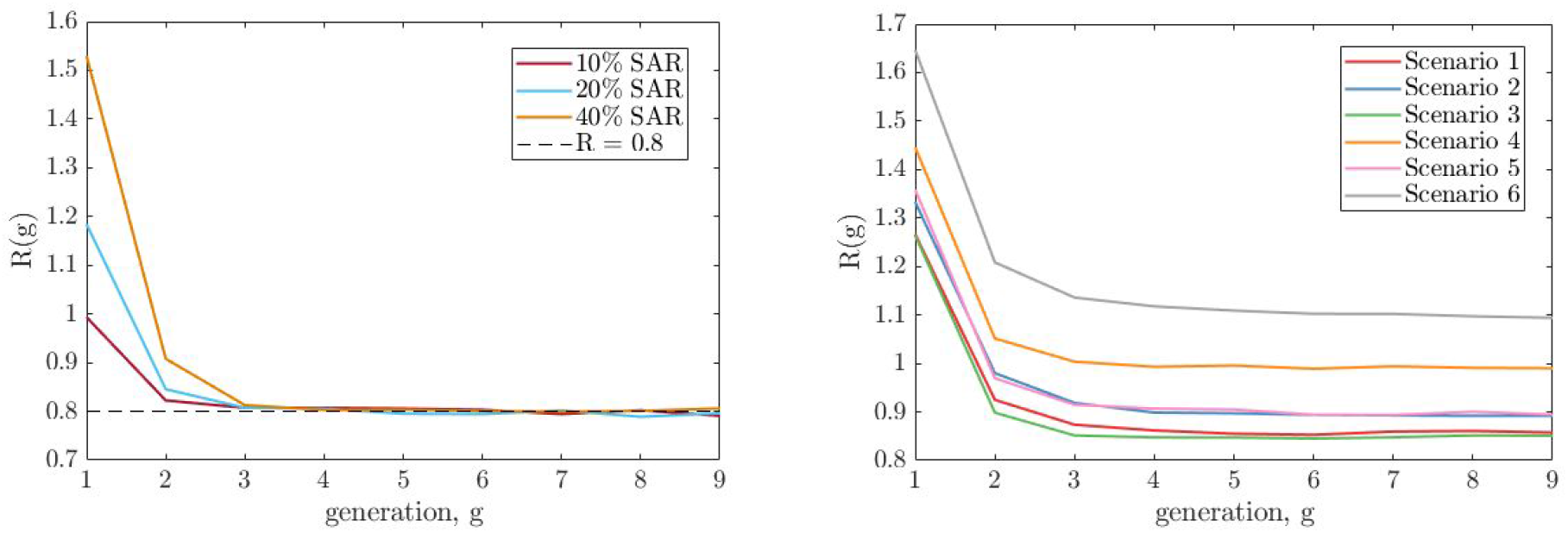
Numerical exploration of *R* by generation. Left shows examples of the method for which ε was fitted to satisfy *R(4)* = 0.8 under our baseline parameters for different values of SAR_HH_. Right shows *R(g)* by generation from each of our scenarios from our baseline assumption. In both plots, *R(g)* decreases over the first few generations, before reaching an equilibrium value which persists over multiple generations.

We also consider the fatality incidence increase caused by various social bubble intervention scenarios. This is calculated as the risk ratio, i.e. the number of deaths given the intervention over the number of deaths given no intervention. Deaths were counted over the fifth to the tenth generation model, and age dependent case fatality ratios were used to project the number of deaths [33].

### Model parameterisation

**Table S3:** Key model parameters and assumptions.

| Parameter | Description | Value (base case) | Value (Sensitivity) | Source |
| --- | --- | --- | --- | --- |
|  | Household structure and age distribution |  |  | [33] |
| $\tau_H$ | Transmission rate for an adult, in a two person household | 0.64 (20% SAR) | 0.29 (10% SAR)<br>1.59 (40% SAR) | Calibrated to SAR [41] |
| $\tau_B$ | Transmission rate for an adult within the bubble | $0.5 \tau_H$ | $1 \tau_H$<br>$0.1 \tau_H$ | assumption |
|  | Relative transmissibility of a child and older adult vs adults | 1 and 1 | 0.64 and 2.9 | [8,53] |
|  | Relative susceptibility of a child and older adult vs adults | 0.5 and 1 | 0.79 and 1.25 | [53] |
|  | Infection fatality rate | In 10y age bands |  | [39] |
| $R_e$ | Net reproduction number | 0.8 | 0.7, 0.9 | [44] |
| $\varepsilon$ | Rate of infection from the community | 1.49<br>(20% SAR) | 1.735<br>(10% SAR)<br>1.15<br>(40% SAR) | Calibrated to $R_e$ given $\tau_H$ |

### Constructing the synthetic population from census data

We used data from the 2011 census of England and Wales (available on request from the Office for National Statistics as dataset CT1088) to construct a distribution of age-stratified household compositions in terms of ten-year age bands. Each household composition consisted of the number of individuals in each age band belonging to the household. We assigned probabilities to each composition observed in the census data based on the frequency of its appearance, and then used these probabilities to construct our simulated household populations. This gave us a synthetic population whose age structure was comparable with that of England and Wales and whose household compositions reflected the observed correlations between the ages of household occupants. In particular, this formulation should realistically capture the generational structure of households in England and Wales, which we expect to be an important factor in transmission across age classes. The data available to us covered households containing six individuals or fewer. Ten-year age band data is not publicly available from the ONS for larger households. Households of size six or less account for 98.2% of the households in England and Wales, and contain 97.8% of their combined population, so that the loss accuracy induced by this cutoff is likely to be minimal.

**Table S4:** Estimated reproduction number for the considered scenarios under the assumption that all eligible households pair up and thereby form exclusive social bubbles, under our base parameterisation.

| # | Scenario description | Reproduction number, assuming $SAR_{HH} =$ | | |
| --- | --- | --- | --- | --- |
|  |  | 10% | **20%** | 40% |
| C1 | Comparator 1 (Baseline):<br>Current lockdown with all households adhering to the lockdown rules | 0.8 | 0.8 | 0.8 |
| C2 | Comparator 2:<br>Unclustered fixed additional contacts, population force of infection matched to Scenario 6 | 0.93 | 1.14 | 1.66 |
| C3 | Comparator 3:<br>Unclustered varying additional contacts, population force of infection matched to Scenario 6 | 0.95 | 1.16 | 1.82 |
| 1 | Allowing all households with primary school age children to pair up | 0.83 | 0.85 | 0.95 |
| 2 | Allowing all households with children of any age to pair up | 0.83 | 0.90 | 1.03 |
| 3 | All single occupancy households to link up with other single occupancy households | 0.84 | 0.85 | 0.88 |
| 4 | All single occupancy households to link up with any other household | 0.90 | 1.00 | 1.11 |
| 5 | Scenarios 1 and 3 | 0.84 | 0.90 | 1.01 |
| 6 | All households pair up | 0.95 | 1.11 | 1.34 |

**Table S5:** Estimated increase in fatalities in generations 5 to 9 for the considered scenarios under the assumption that all eligible households pair up and thereby form exclusive social bubbles and that transmission rates, under our base parameterisation.

| # | Scenario description | Increase in fatalities, assuming $SAR_{HH} =$ | | |
| --- | --- | --- | --- | --- |
|  |  | 10% | 20% | 40% |
| C1 | Comparator 1 (Baseline):<br>Current lockdown with all households adhering to the lockdown rules | 1 | 1 | 1 |
| C2 | Comparator 2:<br>Unclustered fixed additional contacts, population force of infection matched to Scenario 6 | 2.65 | 8.45 | 40.64 |
| C3 | Comparator 3:<br>Unclustered varying additional contacts, population force of infection matched to Scenario 6 | 2.78 | 9.43 | 46.87 |
| 1 | Allowing all households with primary school age children to pair up | 1.18 | 1.49 | 2.57 |
| 2 | Allowing all households with children of any age to pair up | 1.20 | 1.98 | 4.09 |
| 3 | All single occupancy households to link up with other single occupancy households | 1.20 | 1.39 | 1.90 |
| 4 | All single occupancy households to link up with any other household | 2.05 | 3.77 | 6.98 |
| 5 | Scenarios 1 and 3 | 1.39 | 2.24 | 4.14 |
| 6 | All households pair up | 3.13 | 7.82 | 20.26 |

**Supplementary Figure S2:**
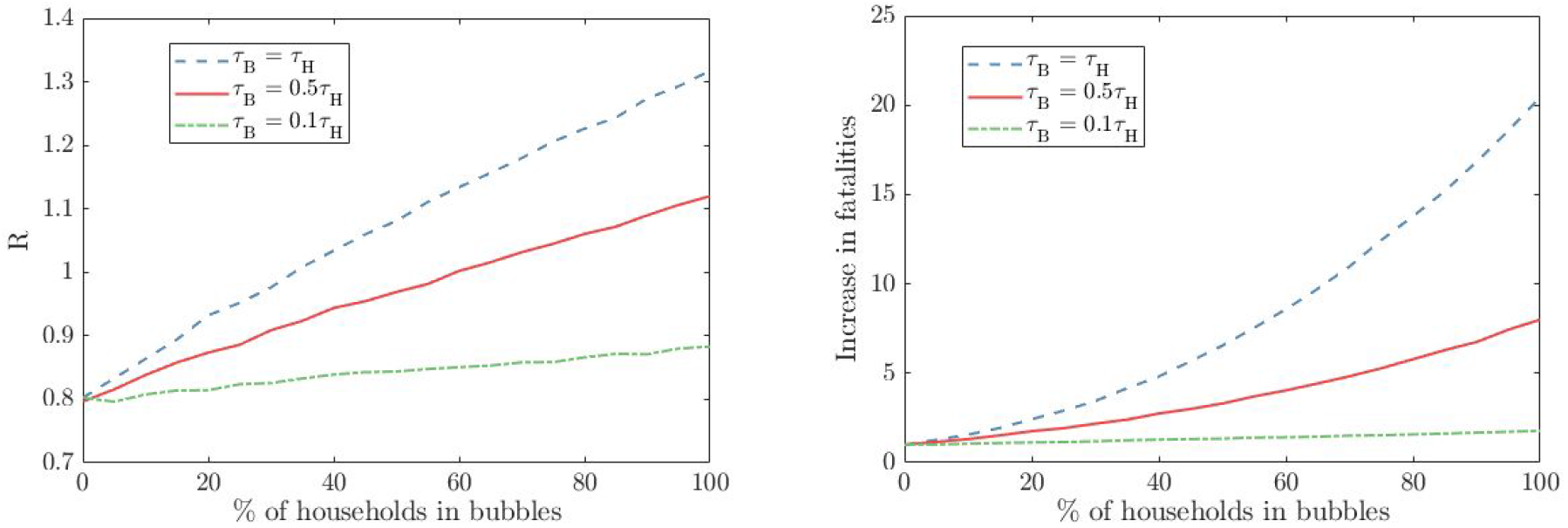
The impact of uptake on *R* and fatality. Here we consider the impact varying levels of uptake has on the reproduction number, *R*, and the fatality incidence ratio. We consider this for our baseline parameters, at varying levels of transmission across bubble contacts (*T_B_* = *T_H_* in blue, *τ_B_* = 0.5 *τ_H_* in red, *τ_B_* = 0.1 *τ_H_* in green). We observe that R scales approximately linearly with uptake, with the gradient of increase dependent on transmission rate across bubble contacts.

**Supplementary Figure S3:**
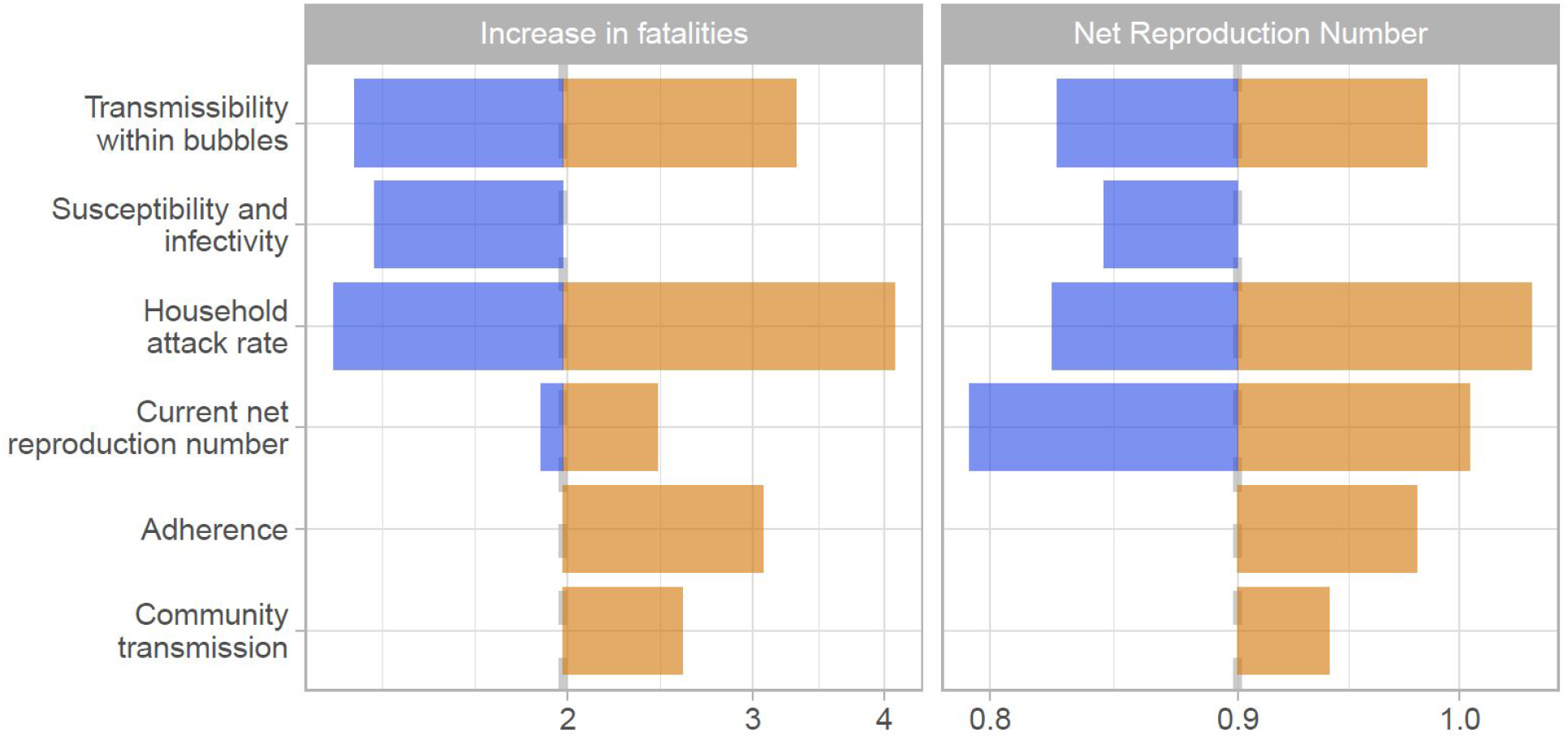
The tornado diagram shows a univariate sensitivity analysis on the expected increase in fatalities and the net reproduction number for scenario 2. The color coding is based on factors determining higher risk (orange) and lower risk (blue) for scenario 1. The base case estimate is indicated through the dashed grey vertical line. The sensitivity scenarios are: transmission across individuals of households sharing a bubble is 90% or 0% lower than that within a household instead of 50%, the relative susceptibility to infection of children and older adults compared to adults is 79% and 125% while the relative transmissibility is 64% and 290%, the secondary attack rate in the household is 10% or 40% instead of 20%, *R_e_* is 0.7 or 0.9 instead of 0.8, that 50% of bubbles do not adhere to the recommendations but pair up to bubbles with four households rather than perfect adherence and that the risk of a household to get infected from the community is proportional to the household size instead of being equal across households.

**Supplementary Figure S4:**
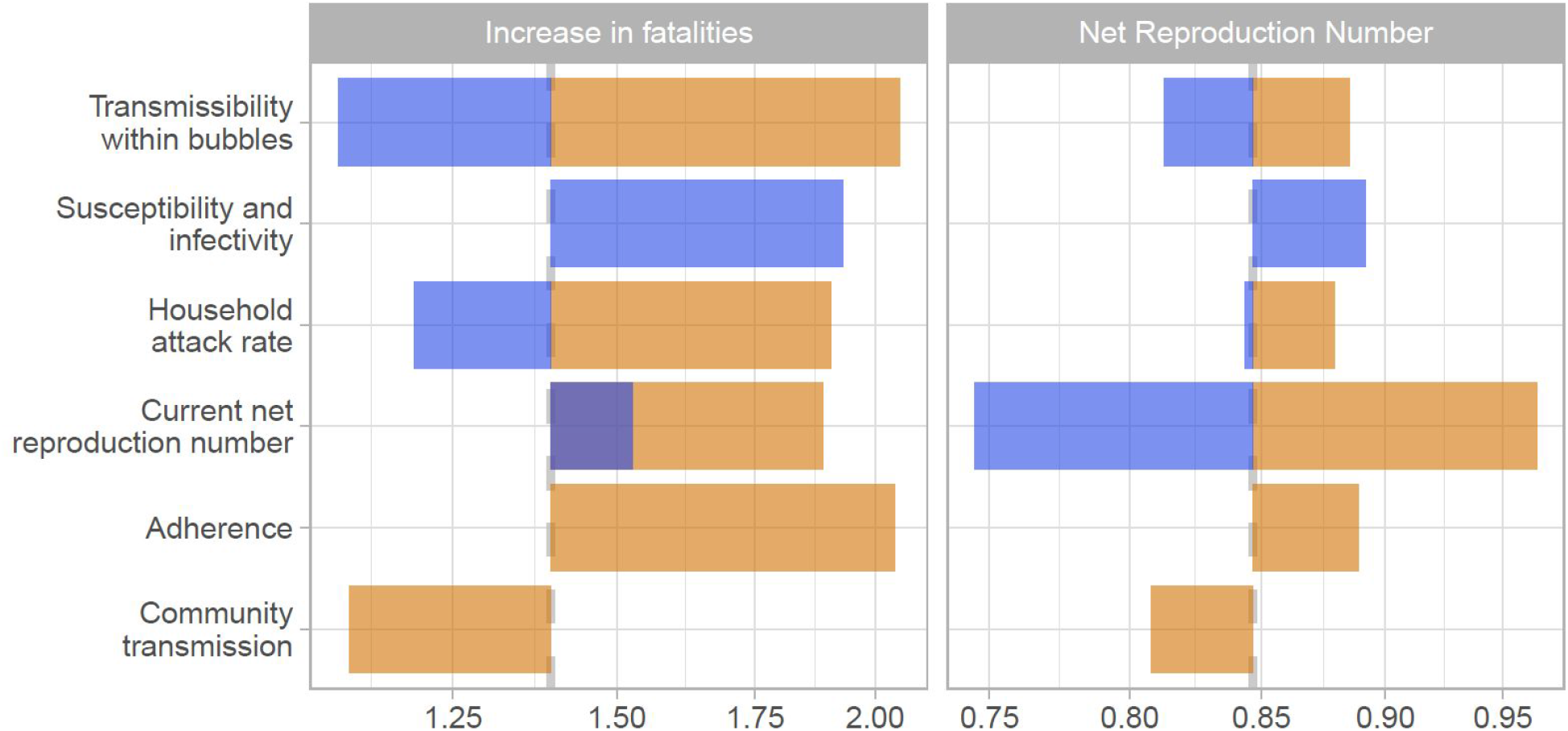
The tornado diagram shows a univariate sensitivity analysis on the expected increase in fatalities and the net reproduction number for scenario 3. The color coding is based on factors determining higher risk (orange) and lower risk (blue) for scenario 1. The base case estimate is indicated through the dashed grey vertical line. The sensitivity scenarios are: transmission across individuals of households sharing a bubble is 90% or 0% lower than that within a household instead of 50%, the relative susceptibility to infection of children and older adults compared to adults is 79% and 125% while the relative transmissibility is 64% and 290%, the secondary attack rate in the household is 10% or 40% instead of 20%, *R_e_* is 0.7 or 0.9 instead of 0.8, that 50% of bubbles do not adhere to the recommendations but pair up to bubbles with four households rather than perfect adherence and that the risk of a household to get infected from the community is proportional to the household size instead of being equal across households.

**Supplementary Figure S5:**
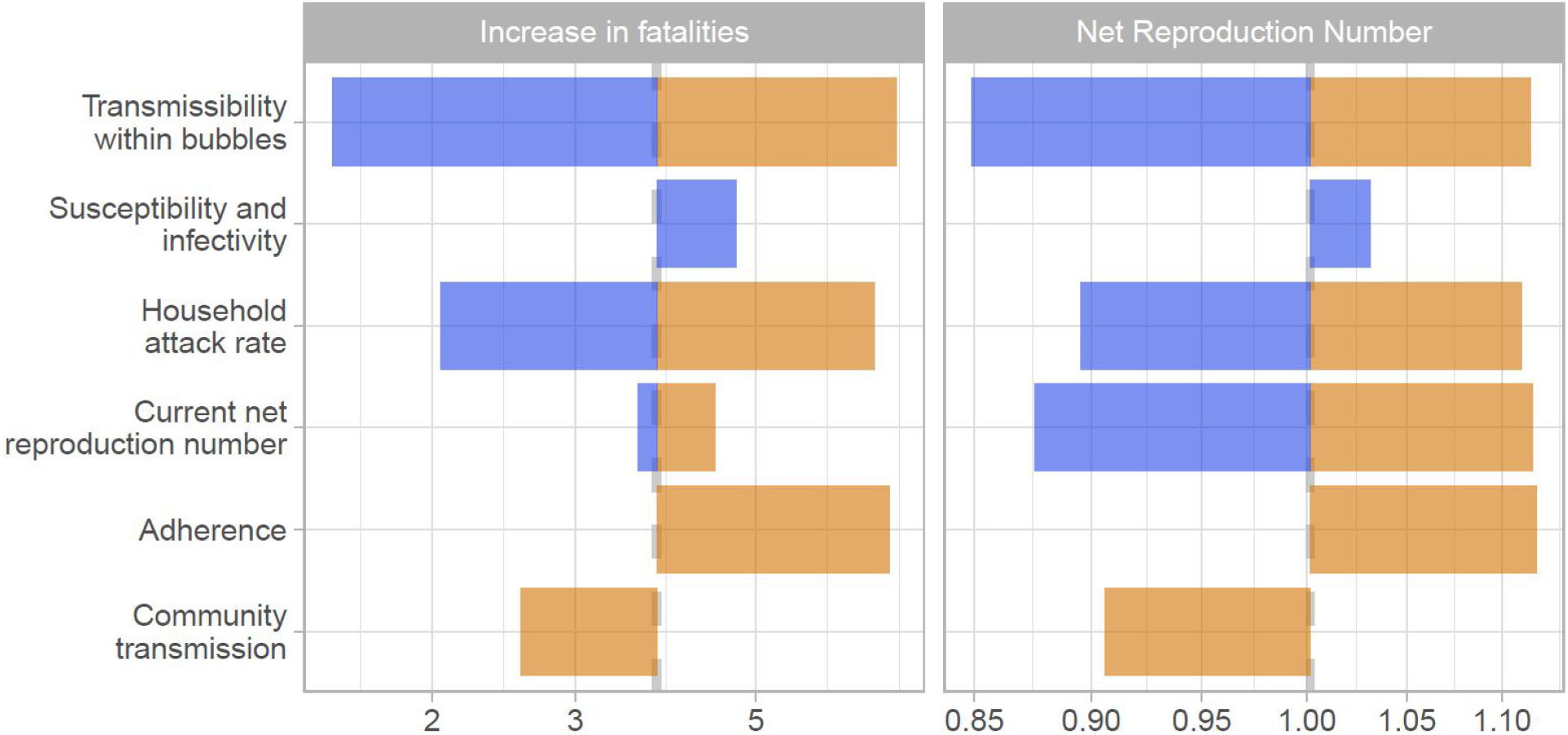
The tornado diagram shows a univariate sensitivity analysis on the expected increase in fatalities and the net reproduction number for scenario 4. The color coding is based on factors determining higher risk (orange) and lower risk (blue) for scenario 1. The base case estimate is indicated through the dashed grey vertical line. The sensitivity scenarios are: transmission across individuals of households sharing a bubble is 90% or 0% lower than that within a household instead of 50%, the relative susceptibility to infection of children and older adults compared to adults is 79% and 125% while the relative transmissibility is 64% and 290%, the secondary attack rate in the household is 10% or 40% instead of 20%, *R_e_* is 0.7 or 0.9 instead of 0.8, that 50% of bubbles do not adhere to the recommendations but pair up to bubbles with four households rather than perfect adherence and that the risk of a household to get infected from the community is proportional to the household size instead of being equal across households.

**Supplementary Figure S6:**
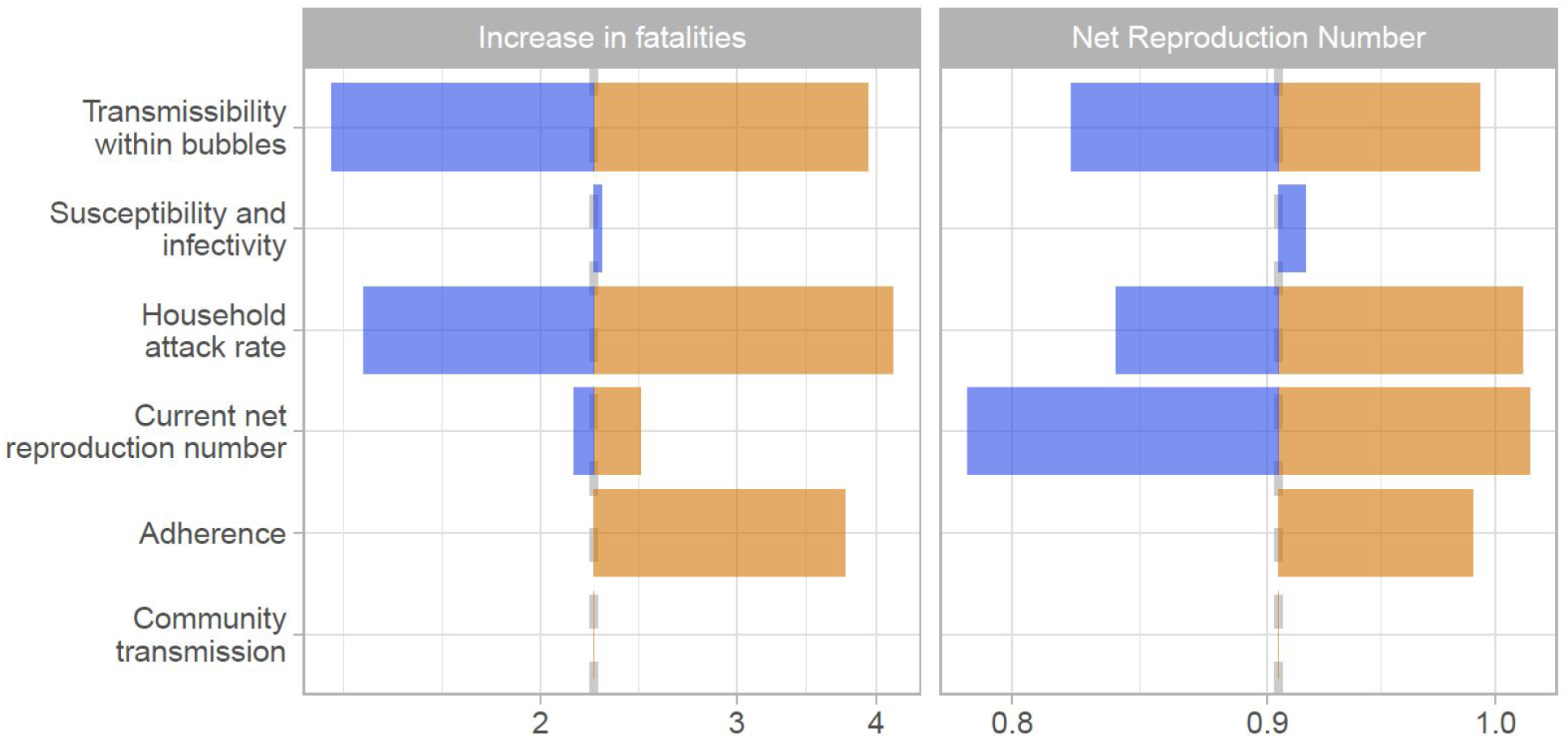
The tornado diagram shows a univariate sensitivity analysis on the expected increase in fatalities and the net reproduction number for scenario 5. The color coding is based on factors determining higher risk (orange) and lower risk (blue) for scenario 1. The base case estimate is indicated through the dashed grey vertical line. The sensitivity scenarios are: transmission across individuals of households sharing a bubble is 90% or 0% lower than that within a household instead of 50%, the relative susceptibility to infection of children and older adults compared to adults is 79% and 125% while the relative transmissibility is 64% and 290%, the secondary attack rate in the household is 10% or 40% instead of 20%, *R_e_* is 0.7 or 0.9 instead of 0.8, that 50% of bubbles do not adhere to the recommendations but pair up to bubbles with four households rather than perfect adherence and that the risk of a household to get infected from the community is proportional to the household size instead of being equal across households.

**Supplementary Figure S7:**
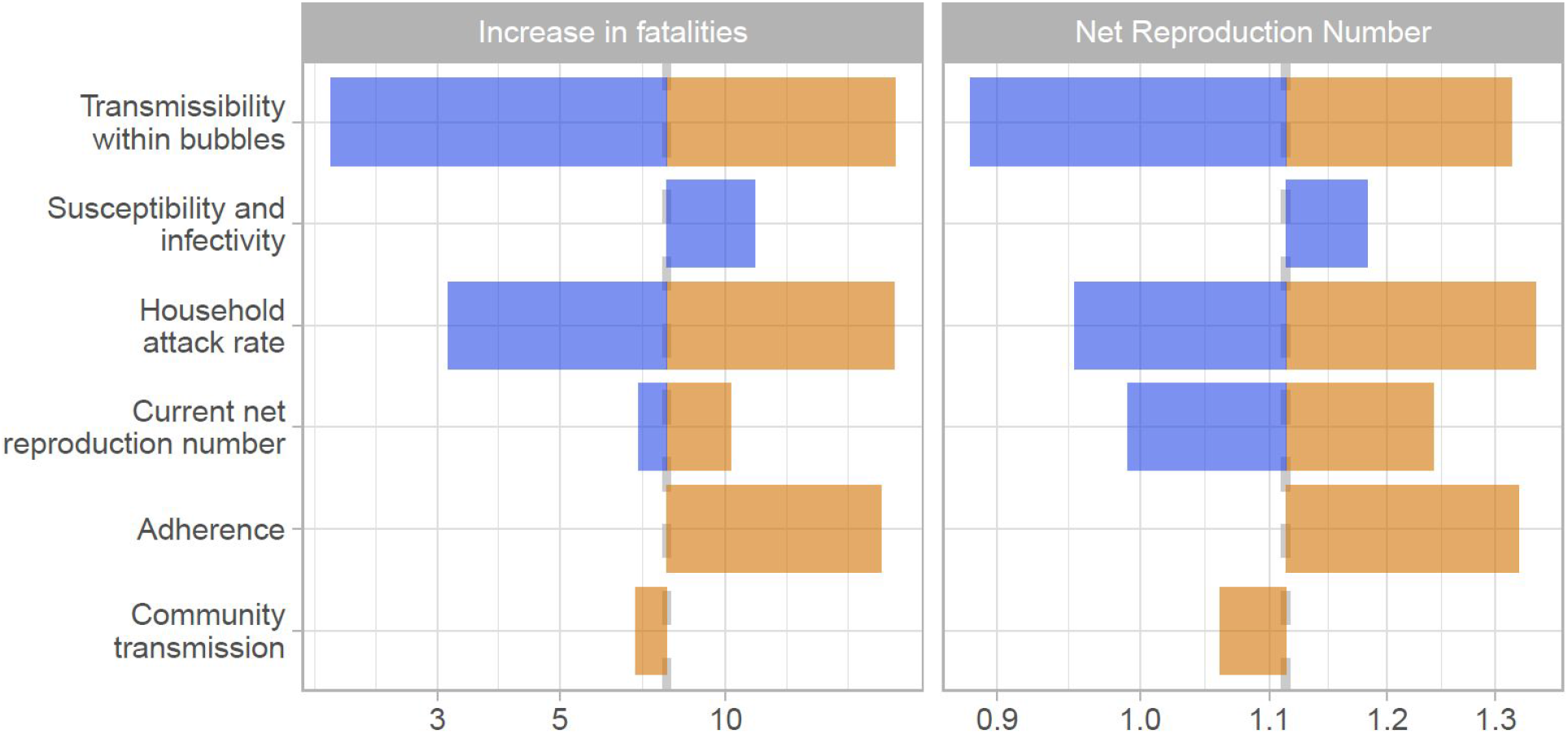
The tornado diagram shows a univariate sensitivity analysis on the expected increase in fatalities and the net reproduction number for scenario 6. The color coding is based on factors determining higher risk (orange) and lower risk (blue) for scenario 1. The base case estimate is indicated through the dashed grey vertical line. The sensitivity scenarios are: transmission across individuals of households sharing a bubble is 90% or 0% lower than that within a household instead of 50%, the relative susceptibility to infection of children and older adults compared to adults is 79% and 125% while the relative transmissibility is 64% and 290%, the secondary attack rate in the household is 10% or 40% instead of 20%, *R_e_* is 0.7 or 0.9 instead of 0.8, that 50% of bubbles do not adhere to the recommendations but pair up to bubbles with four households rather than perfect adherence and that the risk of a household to get infected from the community is proportional to the household size instead of being equal across households.

**Supplementary Figure S8:**
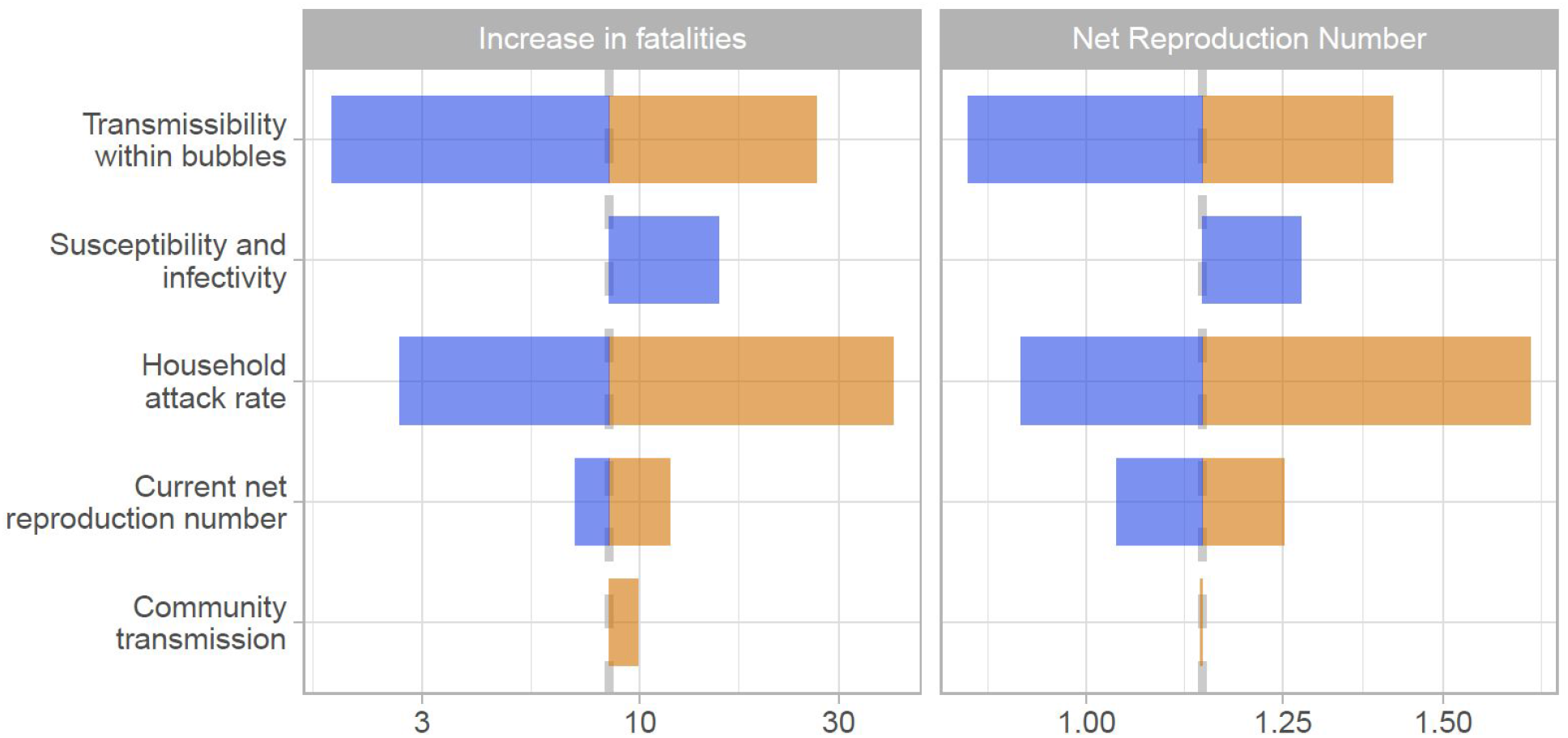
The tornado diagram shows a univariate sensitivity analysis on the expected increase in fatalities and the net reproduction number for scenario C2. The color coding is based on factors determining higher risk (orange) and lower risk (blue) for scenario 1. The base case estimate is indicated through the dashed grey vertical line. The sensitivity scenarios are: transmission across individuals of households sharing a bubble is 90% or 0% lower than that within a household instead of 50%, the relative susceptibility to infection of children and older adults compared to adults is 79% and 125% while the relative transmissibility is 64% and 290%, the secondary attack rate in the household is 10% or 40% instead of 20%, *R_e_* is 0.7 or 0.9 instead of 0.8, that 50% of bubbles do not adhere to the recommendations but pair up to bubbles with four households rather than perfect adherence and that the risk of a household to get infected from the community is proportional to the household size instead of being equal across households.

**Supplementary Figure S9:**
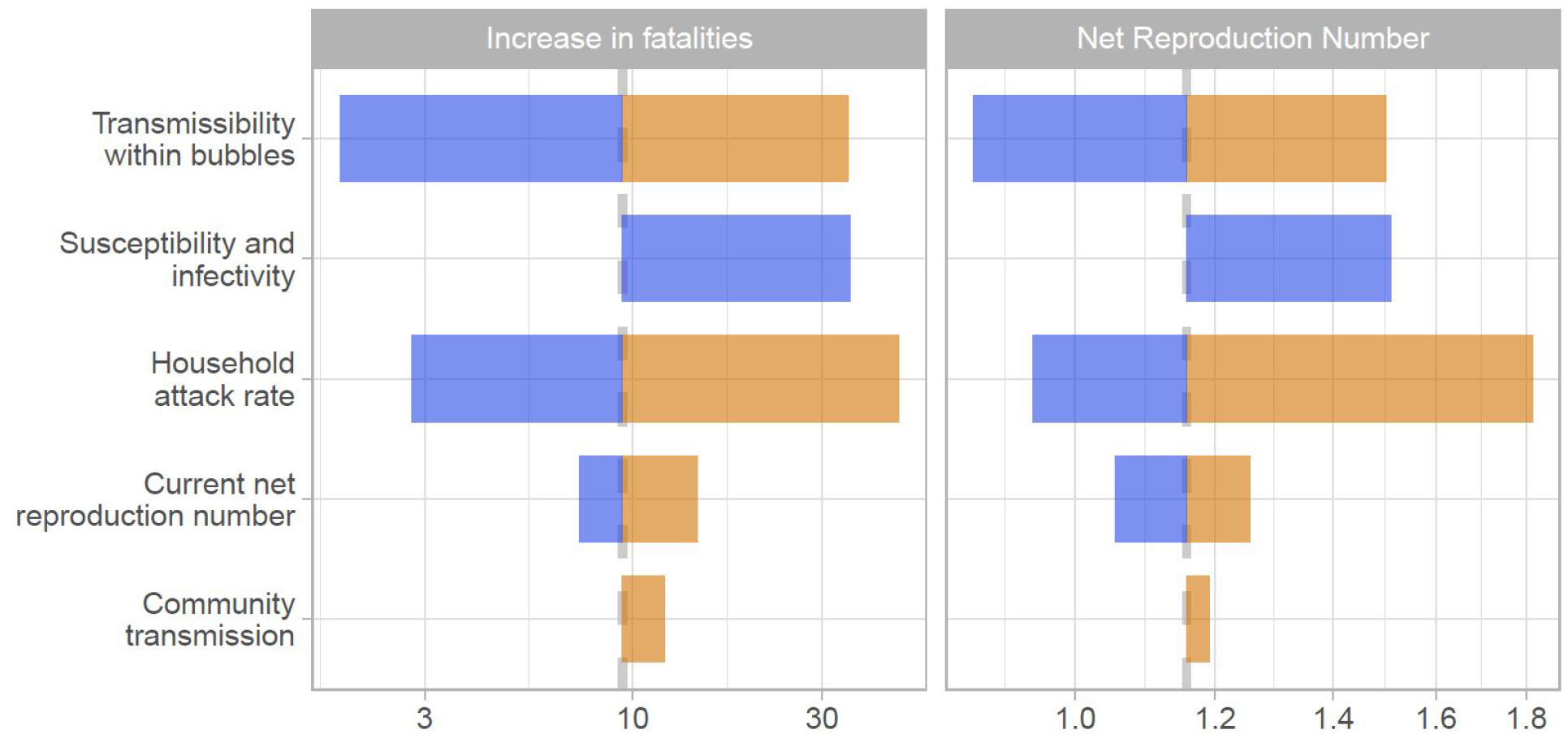
The tornado diagram shows a univariate sensitivity analysis on the expected increase in fatalities and the net reproduction number for scenario C3. The color coding is based on factors determining higher risk (orange) and lower risk (blue) for scenario 1. The base case estimate is indicated through the dashed grey vertical line. The sensitivity scenarios are: transmission across individuals of households sharing a bubble is 90% or 0% lower than that within a household instead of 50%, the relative susceptibility to infection of children and older adults compared to adults is 79% and 125% while the relative transmissibility is 64% and 290%, the secondary attack rate in the household is 10% or 40% instead of 20%, *R_e_* is 0.7 or 0.9 instead of 0.8, that 50% of bubbles do not adhere to the recommendations but pair up to bubbles with four households rather than perfect adherence and that the risk of a household to get infected from the community is proportional to the household size instead of being equal across households.

## References

1 Flaxman S, Mishra S, Gandy A, et al. Report 13: Estimating the number of infections and the impact of non-pharmaceutical interventions on COVID-19 in 11 European countries. Imperial College London, 2020 DOI:10.25561/77731.

2 Jit M, Jombart T, Nightingale ES, et al. Estimating number of cases and spread of coronavirus disease (COVID-19) using critical care admissions, United Kingdom, February to March 2020 Eurosurveillance 2020; 25: 2000632.

3 Cluver L, Lachman JM, Sherr L, et al. Parenting in a time of COVID-19. Lancet Lond Engl 2020; 395: e64.

4 Forman R, Atun R, McKee M, Mossialos E. 12 Lessons Learned from the Management of the Coronavirus Pandemic. Health Policy Amst Neth 2020; published online May 15. DOI:10.1016/j.healthpol.2020.05.008.

5 Appleby J. Tackling covid-19: are the costs worth the benefits? BMJ 2020; 369. DOI:10.1136/bmj.m1496.

6 Staniscuaski F, Reichert F, Werneck FP, et al. Impact of COVID-19 on academic mothers. Science 2020; 368: 724–724.

7 Gilbert M, Dewatripont M, Muraille E, Platteau J-P, Goldman M. Preparing for a responsible lockdown exit strategy. Nat Med 2020; 26: 643–4.

8 Keeling MJ, Hill E, Gorsich E, et al. Predictions of COVID-19 dynamics in the UK: short-term forecasting and analysis of potential exit strategies. medRxiv 2020;: 2020.05.10.20083683.

9 Davies NG, Kucharski AJ, Eggo RM, Gimma A, Group CC-19 W, Edmunds WJ. The effect of non-pharmaceutical interventions on COVID-19 cases, deaths and demand for hospital services in the UK: a modelling study. medRxiv 2020;: 2020.04.01.20049908.

10 Coronavirus (COVID-19): framework for decision making-further information-gov-scot. https://www.gov.scot/publications/coronavirus-covid-19-framework-decision-making-further-information/ (accessed May 30, 2020).

11 Ferguson N, Laydon D, Nedjati Gilani G, et al. Report 9: Impact of non-pharmaceutical interventions (NPIs) to reduce COVID19 mortality and healthcare demand. Imperial College London, 2020 DOI:10.25561/77482.

12 Ferretti L, Wymant C, Kendall M, et al. Quantifying SARS-CoV-2 transmission suggests epidemic control with digital contact tracing. Science 2020; 368. DOI:10.1126/science.abb6936.

13 Kucharski AJ, Klepac P, Conlan A, et al. Effectiveness of isolation, testing, contact tracing and physical distancing on reducing transmission of SARS-CoV-2 in different settings. medRxiv 2020;: 2020.04.23.20077024.

14 He X, Lau EHY, Wu P, et al. Temporal dynamics in viral shedding and transmissibility of COVID-19. Nat Med 2020; 26: 672–5.

15 Liu Y, Centre for Mathematical Modelling of Infectious Diseases nCoV Working Group, Funk S, Flasche S. The contribution of pre-symptomatic infection to the transmission dynamics of COVID-2019. Wellcome Open Res 2020; 5: 58.

16 Peto J, Alwan NA, Godfrey KM, et al. Universal weekly testing as the UK COVID-19 lockdown exit strategy. Lancet Lond Engl 2020; 395: 1420–1.

17 Clifford S, Pearson CAB, Klepac P, et al. Effectiveness of interventions targeting air travellers for delaying local outbreaks of SARS-CoV-2. J Travel Med DOI:10.1093/jtm/taaa068.

18 Kraemer MUG, Yang C-H, Gutierrez B, et al. The effect of human mobility and control measures on the COVID-19 epidemic in China. Science 2020; 368: 493–7.

19 Clase CM, Fu EL, Joseph M, et al. Cloth Masks May Prevent Transmission of COVID-19: An Evidence-Based, Risk-Based Approach. Ann Intern Med2020 published online May 22. DOI:10.7326/M20-2567.

20 Lustig S, Biswakarma JJH, Rana D, et al. Effectiveness of Common Fabrics to Block Aqueous Aerosols of Virus-like Nanoparticles. ACS Nano 2020; published online May 21. DOI:10.1021/acsnano.0c03972.

21 Worby CJ, Chang H-H. Face mask use in the general population and optimal resource allocation during the COVID-19 pandemic. medRxiv 2020;: 2020.04.04.20052696.

22 Liang M, Gao L, Cheng C, et al. Efficacy of face mask in preventing respiratory virus transmission: a systematic review and meta-analysis. medRxiv 2020;: 2020.04.03.20051649.

23 Cheetham J. The ‘social bubble’ approach to lockdown easing. BBC News. 2020; published online April 27. https://www.bbc.com/news/world-52424709 (accessed May 12, 2020).

24 Hinsliff G. Social distancing isn’t going to end soon. So how do we live with it? | Gaby Hinsliff. The Guardian. 2020; published online April 23. https://www.theguardian.com/commentisfree/2020/apr/23/social-distancing-social-pods-coronavirus-lockdown (accessed May 12, 2020).

25 Jones A. Two households will be able to meet up in the garden under new Government plans. The Telegraph. 2020; published online May 26.https://www.telegraph.co.uk/politics/2020/05/26/two-households-will-able-meet-garden-new-government-plans/ (accessed May 28, 2020).

26 Flasche S. Of hairclips and coronavirus-how contact clustering may allow a partial lockdown exit for young kids. LSHTM. https://www.lshtm.ac.uk/newsevents/expert-opinion/hairclips-and-coronavirus-how-contact-clustering-may-allow-partial (accessed May 28, 2020).

27 Block P, Hoffman M, Raabe IJ, et al. Social network-based distancing strategies to flatten the COVID-19 curve in a post-lockdown world. Nat Hum Behav 2020;: 1–9.

28 Our Plan to Rebuild: The UK Government’s COVID-19 recovery strategy.;: 31.

29 Staying alert and safe (social distancing). GOV.UK. https://www.gov.uk/government/publications/staying-alert-and-safe-social-distancing/staying-alert-and-safe-social-distancing (accessed May 30, 2020).

30 Minello A. The pandemic and the female academic. Nature 2020; published online April 17. DOI:10.1038/d41586-020-01135-9.

31 Living alone in the coronavirus crisis | Healthwatch Hillingdon. https://healthwatchhillingdon.org.uk/Blog-COVID19-Isolation (accessed May 28, 2020).

32 Feys F, Brokken S, Peuter SD. Risk-benefit and cost-utility analysis for COVID-19 lockdown in Belgium: the impact on mental health and wellbeing. 2020; published online May 22. DOI:10.31234/osf.io/xczb3.

33 2011 Census-Office for National Statistics. https://www.ons.gov.uk/census/2011census (accessed May 28, 2020).

34 Streeck H, Schulte B, Kuemmerer B, et al. Infection fatality rate of SARS-CoV-2 infection in a German community with a super-spreading event. Infectious Diseases (except HIV/AIDS), 2020 DOI:10.1101/2020.05.04.20090076.

35 Held L. A discussion and reanalysis of the results reported in Jones et al. (2020). 2020; published online May 6. DOI:None.

36 Heavey L, Casey G, Kelly C, Kelly D, McDarby G. No evidence of secondary transmission of COVID-19 from children attending school in Ireland, 2020. Eurosurveillance 2020; 25: 2000903.

37 Cauchemez S, Carrat F, Viboud C, Valleron AJ, Boëlle PY. A Bayesian MCMC approach to study transmission of influenza: application to household longitudinal data. Stat Med 2004; 23: 3469–87.

38 Simpson REH. Infectiousness of communicable diseases in the household (measles, chickenpox, and mumps). Lancet Lond Engl 1952; 2: 549–54.

39 Pellis L, Cauchemez S, Ferguson NM, Fraser C. Systematic selection between age and household structure for models aimed at emerging epidemic predictions. Nat Commun 2020; 11: 906.

40 Pellis L, Ball F, Trapman P. Reproduction numbers for epidemic models with households and other social structures. I. Definition and calculation of R0. Math Biosci 2012; 235: 85–97.

41 Verity R, Okell LC, Dorigatti I, et al. Estimates of the severity of coronavirus disease 2019: a model-based analysis. Lancet Infect Dis 2020; 0. DOI:10.1016/S1473-3099(20)30243-7.

42 Meyerowitz-Katz G, Merone L. A systematic review and meta-analysis of published research data on COVID-19 infection-fatality rates. medRxiv 2020;: 2020.05.03.20089854.

43 Bi Q, Wu Y, Mei S, et al. Epidemiology and transmission of COVID-19 in 391 cases and 1286 of their close contacts in Shenzhen, China: a retrospective cohort study. Lancet Infect Dis2020: S1473309920302875.

44 Koh WC, Naing L, Rosledzana MA, et al. What do we know about SARS-CoV-2 transmission? A systematic review and meta-analysis of the secondary attack rate, serial interval, and asymptomatic infection. Infectious Diseases (except HIV/AIDS), 2020 DOI:10.1101/2020.05.21.20108746.

45 Chau NVV, Lam VT, Dung NT, et al. The natural history and transmission potential of asymptomatic SARS-CoV-2 infection. Infectious Diseases (except HIV/AIDS), 2020 DOI:10.1101/2020.04.27.20082347.

46 Jarvis CI, Van Zandvoort K, Gimma A, et al. Quantifying the impact of physical distance measures on the transmission of COVID-19 in the UK. BMC Med 2020; 18: 124.

47 The R number in the UK. GOV.UK. https://www.gov.uk/guidance/the-r-number-in-the-uk (accessed May 26, 2020).

48 MATLAB. Natick, Massachusetts: The MathWorks Inc., 2020.

49 R Core Team. R: A Language and Environment for Statistical Computing. Vienna, Austria: R Foundation for Statistical Computing, 2019 https://www.R-project.org/.

50 Keeling MJ, Tildesley M, Atkins B, et al. The impact of school reopening on the spread of COVID-19 in England. https://warwick.ac.uk/fac/cross_fac/zeeman_institute/zeeman_research/epidemiology/humams/covid19/warwickcovid19modelschoolreopening.pdf (accessed June 5, 2020).

51 Abbott S, Hellewell J, Thompson RN, et al. Estimating the time-varying reproduction number of SARS-CoV-2 using national and subnational case counts. Wellcome Open Res 2020; 5: 112.

52 Kursschwenk bei Kinderbetreuung: Söder greift Grünen-Idee auf. BR24. 2020; published online April 28. https://www.br.de/nachrichten/bayern/kursschwenk-bei-kinderbetreuung-soeder-greift-gruenen-idee-auf,RxSPHcK (accessed June 1, 2020).

53 New Zealand Governtment. COVID-19 Alert System. Unite COVID-19. https://covid19.govt.nz/alert-system/covid-19-alert-system/ (accessed June 1, 2020).

54 Long NJ, Aikman PJ, Appleton NS, et al. Living in bubbles during the coronavirus pandemic: insights from New Zealand. 2020; published online May 13. http://www.lse.ac.uk/ (accessed May 31, 2020).

55 Endo A, Centre for the Mathematical Modelling of Infectious Diseases COVID-19 Working Group, Abbott S, Kucharski AJ, Funk S. Estimating the overdispersion in COVID-19 transmission using outbreak sizes outside China. Wellcome Open Res 2020; 5: 67.

56 Riou J, Althaus CL. Pattern of early human-to-human transmission of Wuhan 2019 novel coronavirus (2019-nCoV), December 2019 to January 2020 Eurosurveillance 2020; 25. DOI:10.2807/1560-7917.ES.2020.25.4.2000058.

57 Diekmann O, Heesterbeek J a. P, Roberts MG. The construction of next-generation matrices for compartmental epidemic models. J R Soc Interface 2010; 7: 873–85.

58 Ball F, Sirl D, Trapman P. Threshold behaviour and final outcome of an epidemic on a random network with household structure. Adv Appl Probab 2009; 41: 765–96.

